# Scaling rules for pandemics: Estimating infected fraction from identified cases for the SARS-Cov-2 Pandemic

**DOI:** 10.1101/2022.09.05.22279599

**Authors:** Mingyang Ma, Mary Zsolway, Ayush Tarafder, Sebastian Doniach, Gyan Bhanot

**Author notes:** Corresponding Author, Address: 136 Frelinghuysen Rd, Busch Campus, Rutgers Univerrsity, Piscataway, NJ 08854, USA.

## Abstract

Using a modified form of the SIR model, we show that, under general conditions, all pandemics exhibit certain scaling rules. Using only daily data for symptomatic, confirmed cases, these scaling rules can be used to estimate: (i) r_eff_, the effective pandemic R-parameter; (ii) f_tot_, the fraction of *exposed* individuals that were infected (symptomatic and asymptomatic); (iii) L_eff,_ the effective latency, the average number of days an infected individual is able to infect others in the pool of susceptible individuals; and (iv) α, the probability of infection per contact between infected and susceptible individuals. We validate the scaling rules using an example and then apply our method to estimate r_eff_, f_tot_, L_eff_ and α for the first phase of the SARS-Cov-2, Covid-19 pandemic for several countries where there was a well separated first peak in identified infected daily cases after the outbreak of the pandemic in early 2020. Our results are general and can be applied to any pandemic.

## INTRODUCTION

A pandemic occurs when a new pathogen enters a naïve population. The recent SARS-Cov-2 pandemic was caused by a Coronavirus, one of a family of large, enveloped, single-stranded RNA viruses that are widespread in animals and usually cause only mild respiratory illnesses in humans [1-5]. In 2003, a new coronavirus emerged, and was named SARS-CoV (Severe Acute Respiratory Syndrome – Corona Virus). This virus caused a life-threatening respiratory disease in humans, with a fatality rate of almost 10% [6,7]. In fact, after an initial burst of interest in development of treatment options, interest in this virus waned. The emergence of the novel coronavirus SARS-CoV-2, identified in December 2019 in Wuhan, China, has since caused a worldwide pandemic [8-13]. SARS-CoV-2 is the seventh known coronavirus to cause pathology in humans [1]. The associated respiratory illness, called COVID-19, ranges in severity from a symptomless infection [8], to common-cold like symptoms, to viral pneumonia, organ failure, neurological complications, and death [9-11]. While the mortality in SARS-CoV-2 infections is lower than in SARS-CoV [9-12], it has more favorable transmission characteristics, a higher reproduction number, a long latency period and an asymptomatic infective phase [13].

The governments of several countries took significant measures to slow the infection rate of Covid-19, such as social distancing, quarantine, identification, tracking and isolation. However, there was no uniform policy, some governments reacted later than others, and some (e.g. Sweden) decided to keep the country open, leaving counter-measures up to individuals. A large amount of consistent public data is now available on the number of tests performed, the number of confirmed infected cases, and the number of deaths in different contexts, such as locations and health conditions [14]. These provide important sources of information for the development and testing of models to estimate pandemic characteristics, guide public policy and assess the efficacy of interventions [15].

It is well known that in most pandemics, confirmed infected cases often seriously underestimate the actual number of infections [16,17]: not everyone who is infected is symptomatic, and not everyone who dies from the disease has been tested [18]. Even the number of reported deaths may be underestimated because of co-mortalities; i.e. COVID-19 increases susceptibility to other diseases and conditions [19]. Moreover, the virus can be transmitted by asymptomatic individuals, who can comprise a substantial portion of the infected population [20], militating against accurate estimates of total infection rates. In this context, as indicated in [21], analytical models can provide useful information.

Dynamical (mechanistic) models have been used for forecasting and for making projections. For example, projections and forecasting models of various types were used as early as February 2020 to determine a reproductive number for SARS-CoV-2 [13]. More generally, multiple research groups have models to estimate Case Fatality Ratios (CFRs) [22], to forecast and project the need for hospital beds [23] and to project and forecast mortality [24]. Among the many applications of models to COVID-19, four variable Susceptible-Exposed-Infective-Recovered (SEIR) models have been used to project the impact of social distancing on mortality [25], three variable Susceptible-Infective-Recovered (SIR) models have been used to estimate case fatality and recovery ratios early in the pandemic [26], and a time delayed SIR has been used to evaluate the effectiveness of suppression strategies [27]. One of the most ambitious dynamical models, which includes 8 state variables, and 16 parameters, was fruitfully applied to evaluate intervention strategies in Italy, in spite of the fact that parameter identifiability could not be assured [28]. There is also some model based evidence that the transmission of the SARS-Cov-2 virus is regulated by temperature and humidity [29]. In this paper, we model the Covid-19 pandemic using an extension of the SIR model [30], which partitions the population into three compartments: Susceptibles (S), Infectious (I) and Removed R. This and other models (using more variables) have been used in a variety of contexts to study the global spread of diseases (For some recent reviews, see [31-33]). The extension of the SIR model developed in this paper differentiates itself from earlier studies in that it provides a way to make an a-posteriori estimate of several useful epidemiological parameters for any pandemic, using only data on confirmed, identified cases.

The question we ask in this paper is the following: Using only daily recorded case data of symptomatic individuals, is it possible to estimate the actual fraction of infected individuals from among the pool of susceptible individuals who contributed to the recorded cases in the region from which the data was collected? We will show that this question can be answered in the affirmative, at least within the context of an extension of the standard epidemiological SIR model [30]. The reason this is possible is that we show that there is a connection between the identified daily cases and the actual number of individuals who remain infected in the population on that day. We also show that in our extended model, this connection leads to general scaling rules for the location of the peak (days from start of the pandemic to the peak in daily cases) and the half width at full maximum in identified daily cases. We will further show that these scaling rules allow an estimate of an “effective” pandemic R-parameter r_eff_, the fraction f_tot_ of exposed individuals who got infected (both symptomatic and asymptomatic), the effective latency L_eff_, the average number of days an infected individual is able to infect others and α, the probability of infection per contact between infected and susceptible individuals. Within our extended model, these results are general and can be applied to any pandemic. After demonstrating the internal consistency of our approach on model data, we apply our method to worldwide daily case data for the first phase of the SARS-Cov-2 (Covid-19) pandemic in 2020 to derive estimates of these parameters for a number of countries where there was a well separated first peak in identified infected daily cases after the outbreak of this pandemic in early 2020

*We note that our results for f*_*tot*_ *represent only the fraction of infected individuals in the “exposed population” in a given region – i*.*e*., *it only applies to the set of susceptible individuals who came into sufficiently close contact with infected individuals for the virus to transmit. This value should not be taken to represent the fraction of infected individuals in the population as a whole, because our analysis does not include those individuals who were sufficiently isolated in some way (e*.*g*., *self-quarantined, wore masks etc*.*), so as to avoid contact with the virus*.

## METHODS: The Extended SIR Model

We assume that each country is a region where a subset of the population consists of interacting individuals who are equally susceptible to infection and once infected, are responsible for virus transmission. The extension we propose is to assume that there are two types of individuals: those who become symptomatic after infection, and those who do not. Daily counts of infected individuals reflect only those who become symptomatic. We also assume that identified symptomatic individuals are no longer able to infect others because, once identified as infected (possibly after confirmatory testing), they would be isolated, confined, or quarantined. On the other hand, asymptomatic individuals, being unaware of their infected state, would continue to infect others until they become non-infective (cured/recovered). We define the start of the pandemic as the day when the number of recorded daily cases begins to rise exponentially towards a well-defined peak (a more practical definition will be provided later), before decreasing to less than half the peak and possibly continuing to decrease further.

Let L_0_ be the average number of days an asymptomatic individual is infective and L_1_ be the average number of days a symptomatic individual is infective. An asymptomatic individual becomes non-infective (recovered/cured) after an average of L_0_ days, while a symptomatic individual would have symptoms on day L_1_ on average and at this point, would be quarantined and unable to infect others. It is reasonable to assume that L_1_ < L_0_. Let 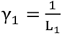 and 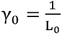 be the rates at which these two types of infected individuals leave the infective pool. Let αbe the probability of infection when an infected individual meets a susceptible (non-infected) individual. Under these assumptions, we can write down a simple extension of the SIR model [1] for the pandemic dynamics as follows:

Let S(t), I(t), R(t) to be the number of Susceptible, Infected and Removed individuals at time t, with S(t) + I(t) + R(t) = N. Here N is the pool of susceptible individuals who were exposed to the virus. At any given time, the I(t) compartment consists of two parts, I_0_(t) and I_1_(t) where the first is a fraction 1-ω of individuals who remain asymptomatic until they recover and the second is a fraction ω of individuals who become symptomatic, are identified and are no longer able to infect others (they move to the “Removed” compartment). The R compartment consists of two types of individuals, a set R_1_(t) derived from I_1_(t), and a set R_0_(t) derived from I_0_(t). The extension of the SIR model that applies in such a situation is defined by the equations:

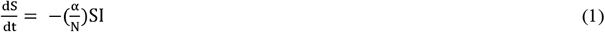

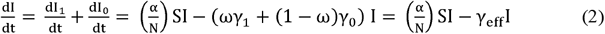

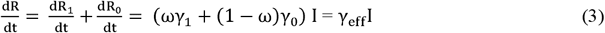

Here

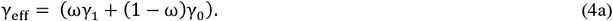

The quantity α is the probability of infection in a single encounter between an infected and susceptible individual. The reciprocal L_eff_ of γ_eff_ is the average effective latency, the average number of days that an infected individual (symptomatic or not) is infective. Thus,

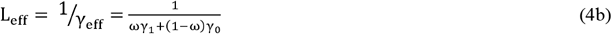

Note that L_eff_ can sometimes be estimated from monitoring and testing of individuals. However, in the general case, it is quite difficult to estimate because its value depends on the fraction of asymptomatic infected cases.

The key quantity in our approach is X(t), the rate at which symptomatic individuals are identified. Thus,

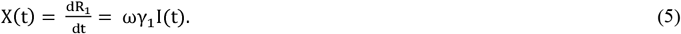

For any pandemic, X(t) is the *observed* daily cases reported from hospitals and testing sites from symptomatic and/or tested individuals. The key observation that leads to the results in this paper is Eq. 5, which asserts that X(t) is proportional to I(t). This proportionality means that the width and location of the peak in X(t) and I(t) are the same.

If we rescale time to 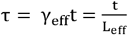 Eq. 1-3 can be rewritten in terms of the fractions s = S/N, i = I/N, r= R/N, r_1_ = R_1_/N and x = X/N as follows:

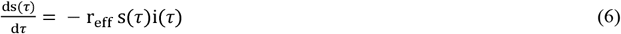

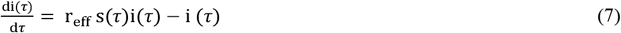

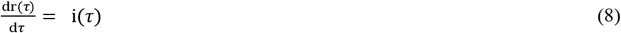

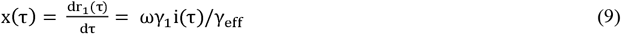

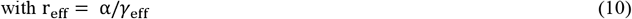

At the start of the pandemic, i.e., at *τ* =0, both i(*τ*) and x(*τ*) are near zero, since a very small fraction of the population is initially infected. It is easy to show that, starting with a small fraction ε of infected cases at *τ* = 0, i(*τ*) and x(*τ*) increase exponentially as 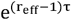 in the interval 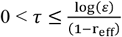 (Appendix A, Eq. A16a,b). Eventually (as we will see in the data and the solution to the model equations), both quantities reach a peak when the fraction of susceptible individuals decreases sufficiently to slow the growth of the pandemic. Finally, i(*τ*) and x(*τ*) diminish to a value near zero when the likelihood of further infections becomes negligible. It is easy to show that for a pandemic to take place at all, r_eff_ must exceed unity. In other words, for r_eff_ ≤1, there is no pandemic and s(∞) =1 (Appendix A (Eq. A8). Thus, r_eff_ is identified as the so called “Pandemic R-parameter”, the single parameter that controls the pandemic dynamics in this model.

To facilitate further discussion, we define the following quantities:

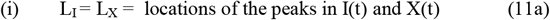

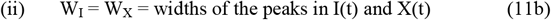

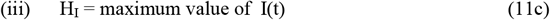

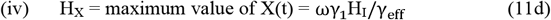

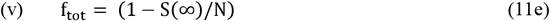

f_tot_ is the total fraction of exposed individuals who become infected, including both symptomatic and asymptomatic cases. This quantity is generally difficult to estimate. However, as noted, we can exploit the fact (Eq. 5 and Eq. 9) that there is a connection between the time dependence of identified symptomatic cases X(t) and x(*τ*), and the time dependence of the total number of cases I(t) and i(*τ*), which includes both symptomatic and asymptomatic cases. Specifically, Eq. 5 says that the location and widths of the peaks in X(t) and I(t) are the same, and Eq. 9 says that location and width of the peaks in x(*τ*) and i(*τ*) are also the same. *The key idea of this paper is that this fact allows one to relate properties of X(t) and I(t) (or x(*_τ_*) and i(*_τ_*)) to estimate r*_*eff*_, *f*_*tot*_, *L*_*eff*_ *and α using only data for X(t)*.

## RESULTS

### I. Universal Scaling Rules for Pandemics

Since time t in physical units (seconds, hours, days) is related to dimensionless time *τ* by 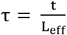, we can relate properties of I(t) and X(t) in Eq. 11 to properties of i(*τ*), and x(*τ*). Thus:

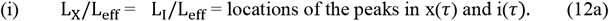

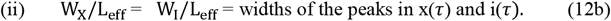

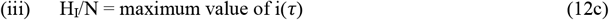

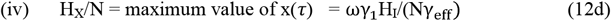

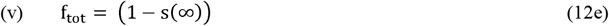

In the limit of large N, it is easy to find an exact formula for H_I_/N (see Eq. A10 and the discussion preceding it in Appendix A):

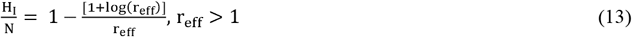

However, although this is interesting, it is not very useful, because relating this quantity to the measurable quantity H_X_/N requires the values of ω, γ_1_ and γ_0_. On the other hand, the relationships in Eq. 12a,b and the fact that all the quantities in Eqs. 11 and 12 are controlled by a single parameter r_eff_ lead to universal scaling rules that can be exploited to estimate f_tot_, r_eff_, L_eff_ and _α_ using only data for X(t). The simplest way to do this is to note that the ratio L_X_/W_X_ is independent of L_eff_ and can be estimated from the measured daily cases X(t). Figures 1a,b show the dependence f_tot_, r_eff_ on L_X_/W_X_ (data in Supplementary Table 1). These results were obtained by numerically solving Eq. 6-8 using the stiff ODE solver ode15s in Matlab for r_eff_ in the range 0.5-6.5. Once r_eff_ (or f_tot_) is known, L_eff_ can be estimated from the functional dependence of L_X_/L_eff_ and W_X_/L_eff_ on these quantities (Figure 1 c-f, data in Supplementary Table 1,).

**Figure 1:**
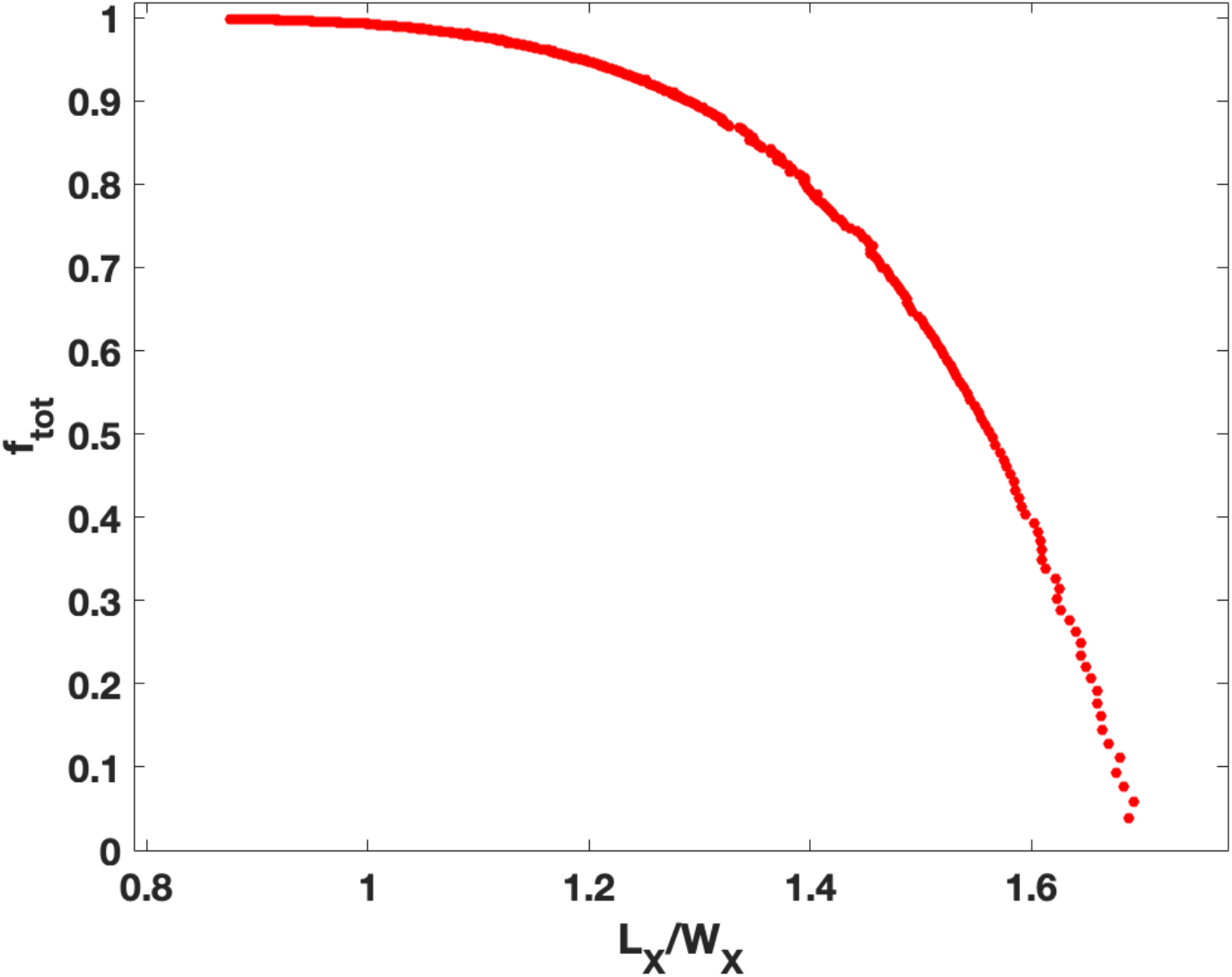
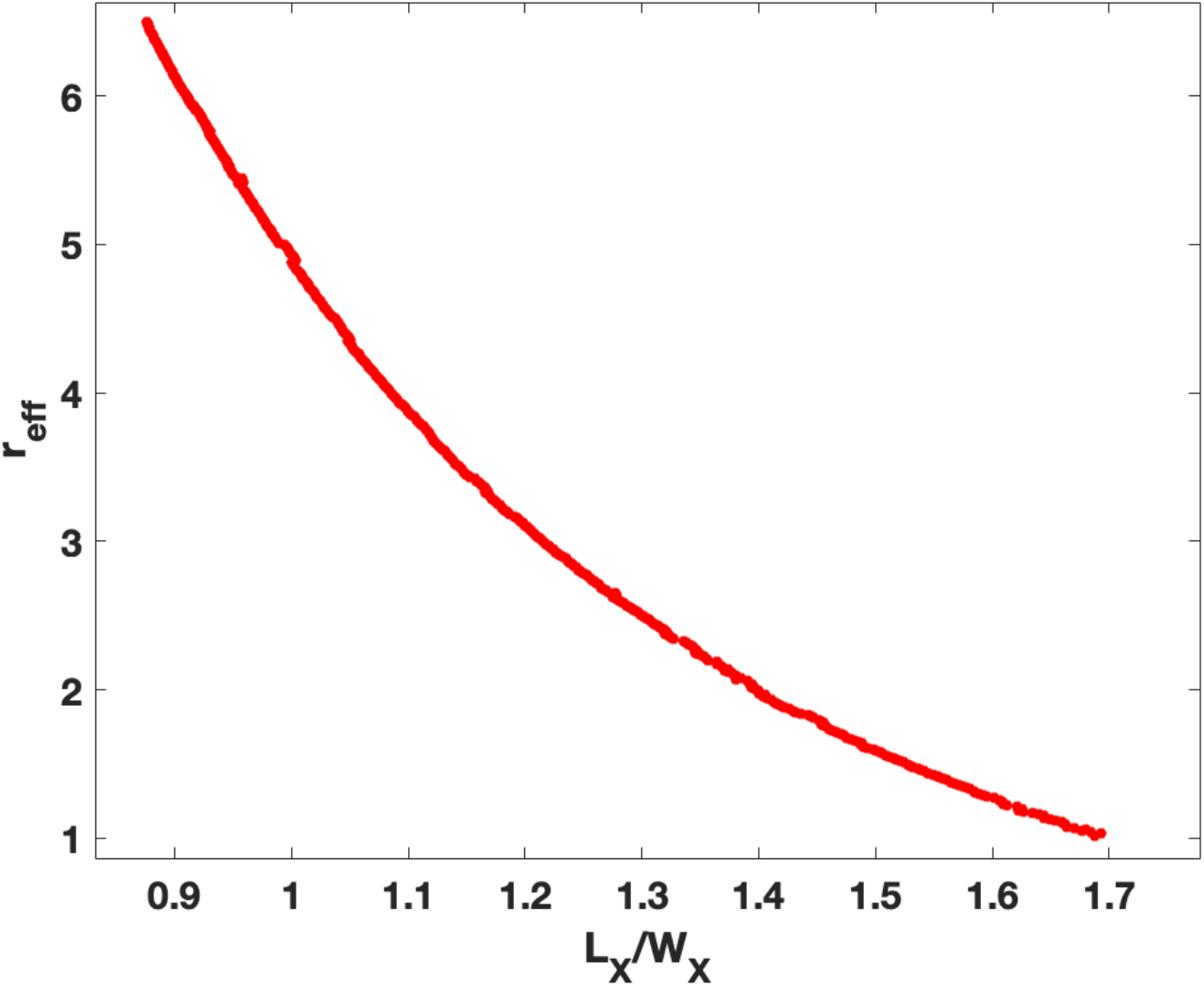
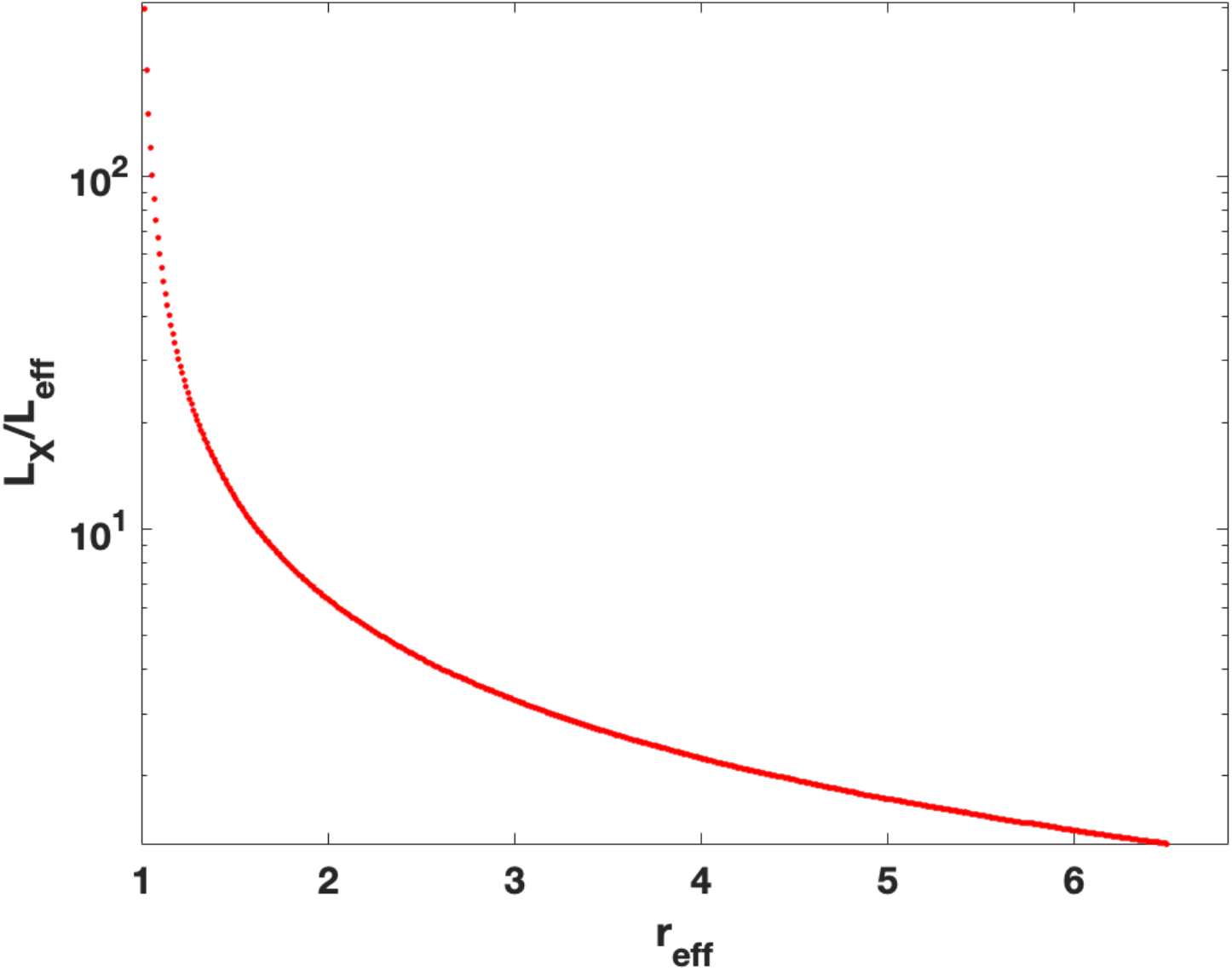
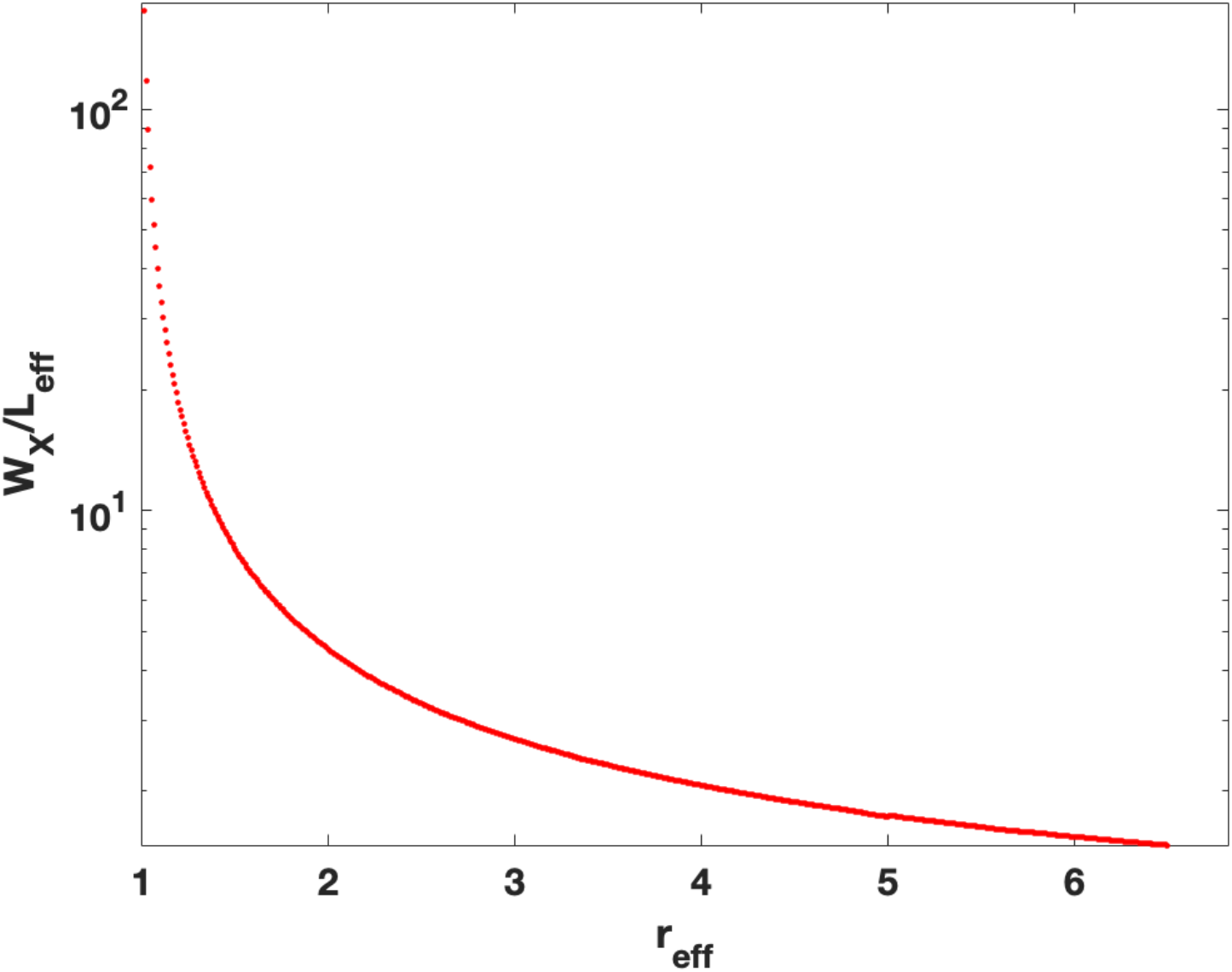
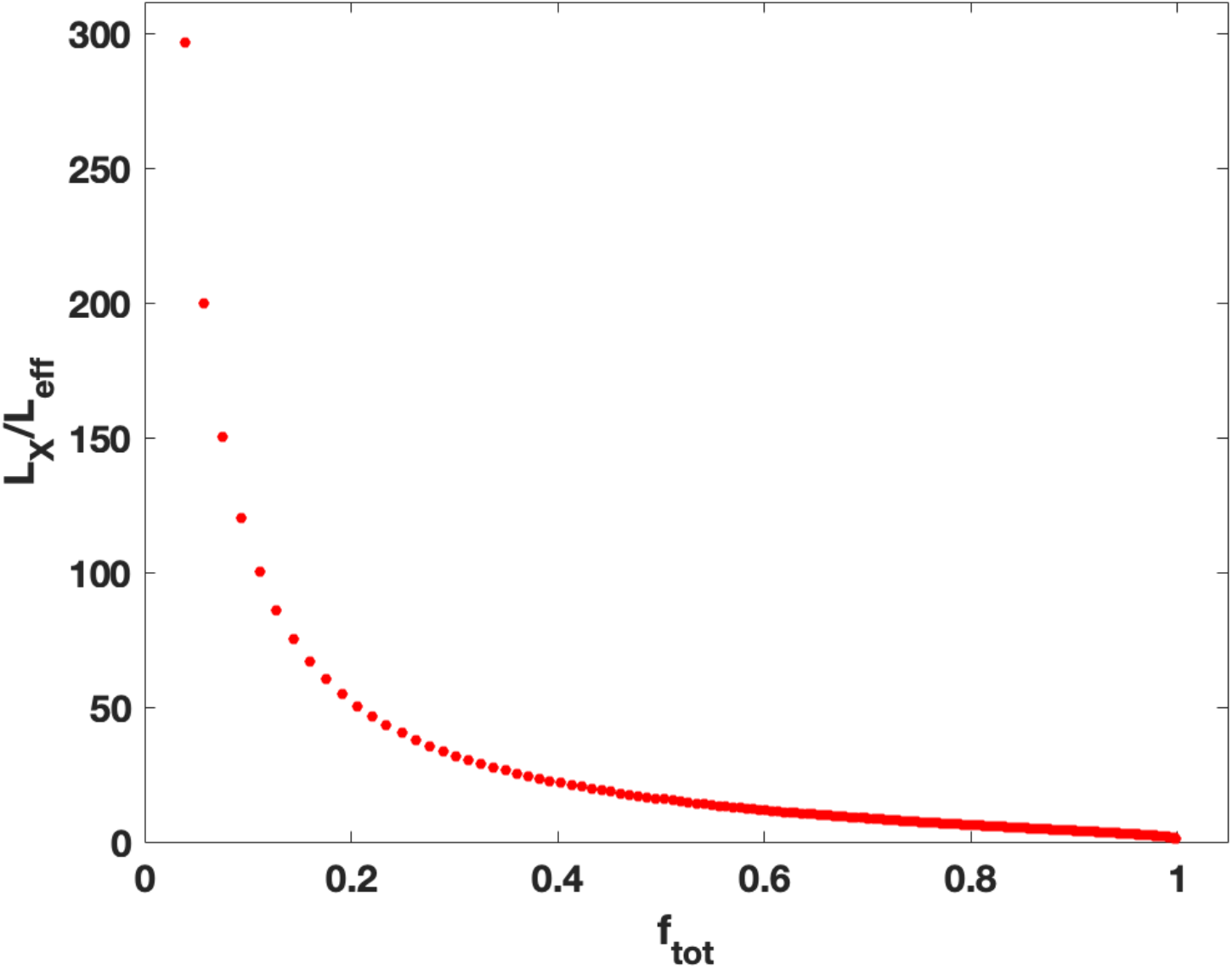
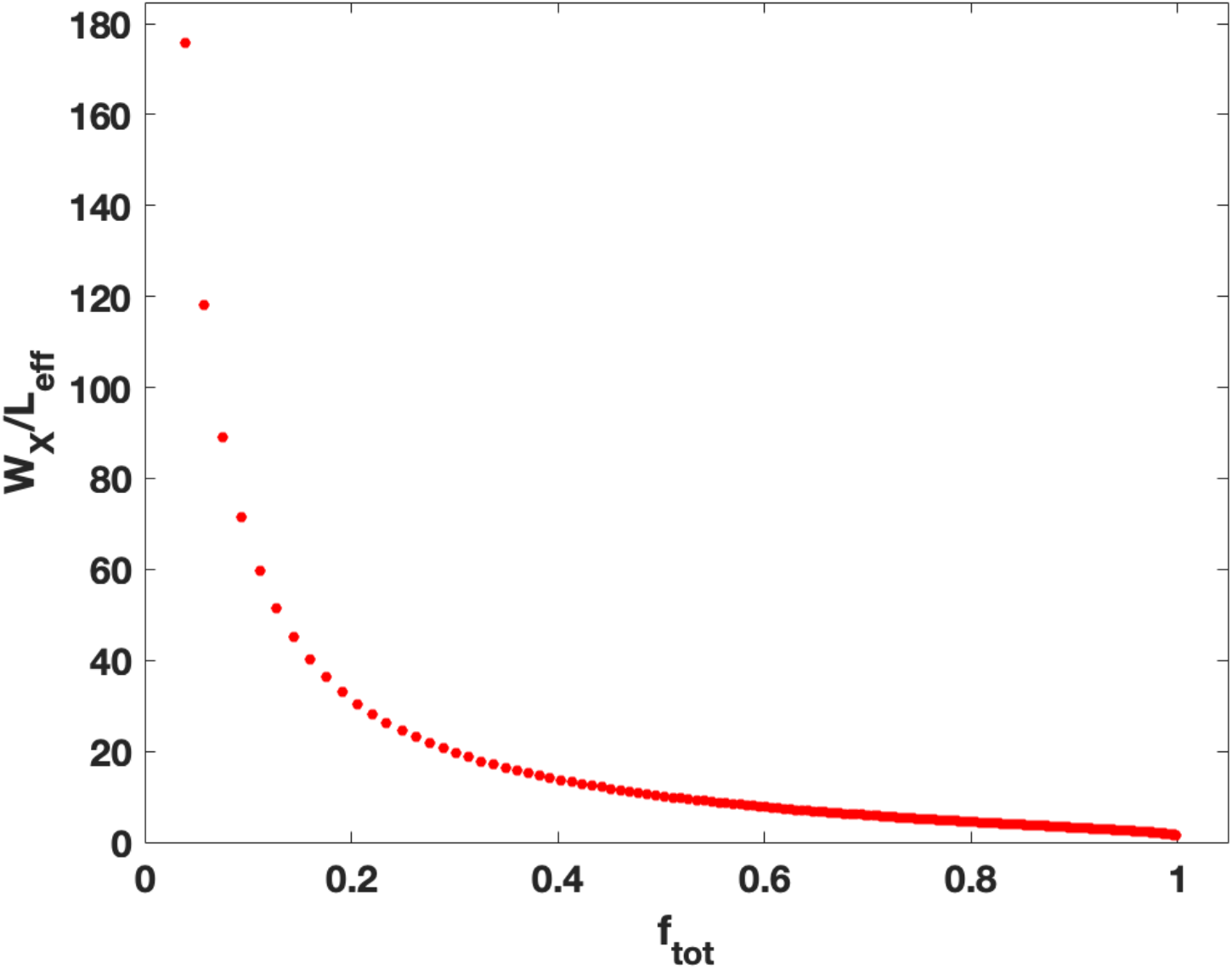
(**a**,**b):** Universal scaling curves in the SIR model for f_fot_ and r_eff_ as functions of the ratio L_X_/W_X_, where L_X_ is the number of days from the start of the pandemic to the location of the peak in daily observed cases X(t) (Eq. 5) and W_X_ is the width of that peak. Note that these functions are independent of L_eff_ and apply to any pandemic. These can be used to find f_tot_ and r_eff_ using the ratio L_X_/W_X_ from data for X(t). (**c-f):** Universal scaling curves in the SIR model for L_X_/L_eff_ and W_X_/L_eff_ as functions of f_fot_ and r_eff_. L_X_ and W_X_ are the location and width of the peak in daily observed cases X(t) (Eq. 5). These data can be used to estimate L_eff_ once r_eff_ and f_tot_ are estimated using Figure 1 (a,b) (data in Supplementary Table 1). Note that these functions are universal and apply to any pandemic.

Figure 1a-f and the data in Supplementary Table 1 are the main results of this paper. Within the limits of the SIR model, these results are universal and apply to any pandemic. For any pandemic, once L_X_ and W_X_ are estimated from data for X(t) in a given region, these data can be used to estimate pandemic parameters.

### II. Inferring f_tot_, r_eff_, L_eff_ and α using only data for X(t)

Appendix B shows an example of the use of the data in Figure 1 and Supplementary Table 1 to estimate f_tot_, r_eff_, and L_eff_ and *α* from L_X_ and W_X_ for one specific set of test parameters used to generate numerical solution of Eq. 1-5. For use in general, we used a minimization procedure that generates initial estimates of f_tot_ and r_eff_ using the experimental value y_e_ = L_X_/W_X_ from the time dependence of X(t). The data in Figure 1b (and Supplementary Table 1) was then used to make an initial estimate r^0^_eff_ for r_eff_ which was iteratively improved by choosing nearby values of r_eff_ to solve Eq. 6-9, compute y(r_eff_) = L_X_(r_eff_) /W_X_(r_eff_) and minimize (y_e_ – y(r_eff_))^2^ as a function of r_eff_.

### III. Application to data for the SARS-CoV-2/Covid-19 pandemic

Worldwide data for confirmed Covid-19 cases and deaths from January 3, 2020 was downloaded from the World Health Organization (WHO) website: https://covid19.who.int/data (Supplementary Table 2). This data estimates the function X(t) in our analysis. Before performing any analysis, the data for daily cases was averaged over eleven days to reduce noise. Averaging over nine, seven or five days did not change the results.

Our model assumes that there was a single circulating strain of the virus that infected a homogeneous set of individuals in a given region who were equally susceptible to infection (uniform immune response). The model also assumes that exposed individuals observed the same rules regarding the use of masks/isolation/quarantine, there was no significant variation in population density among them, little variation in their movements, and equal vaccination status. Symptomatic cases were equally likely to be identified across the region, and consistently obeyed (or disobeyed) rules regarding quarantine, testing, etc. Since we cannot correct for these effects in this paper, we present the results only as proof of concept, and apply our method only to the first wave of the Covid-19 pandemic. For this first wave of the pandemic, the world population was naïve to the virus (no immunity) so that everyone was susceptible. Moreover, at least some of the other assumptions of the model did apply in some countries, such as homogeneity of response, lack of vaccines resulting in no innate immunity, standard medical protocols (and in some cases testing for viral RNA) used in identifying cases, and a single circulating strain of the virus.

We also apply the model only to countries where the data for measured daily cases showed a clear exponential rise from a few cases followed by a clear peak in daily cases with a measurable half width at full maximum for the first phase of the pandemic, which took place in most countries between January 1, 2020, and August 31, 2020. We also require that this initial peak not overlap with subsequent peaks. Thirty-four countries satisfied these conditions. For these, the values of L_X_ and W_X_ were determined for the first peak in daily cases from the data for daily identified cases X(t) (raw data was averaged over 11 days to find X(t)) and f_tot_, r_eff_, L_eff_ and α values were estimated as follows: From the ratio L_X_/W_X_, the value of r_eff_ was inferred by interpolation of the numerical solution data of the scaled model equations (Eq 6-9, Supplementary Table 1). The model solution for x(*τ*) at this value of r_eff_ was then mapped to the data for X(t) by matching the location and heights of the peaks in x(*τ*) and X(t) and rescaling the *τ* axis to the t axis to match the half width of the data for X(t). Since 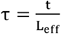, this determines L_eff_ from the rescaling *τ* → t.

Errors in the parameters were determined by varying L_X_ and W_X_ by +/- 1 and recomputing them as described above. To identify the “start” date of the pandemic, which affects the estimate of L_X_, we used the procedure described in Appendix B and which was also used in generating the data in Supplementary Table 1: The start date was chosen as the day when the measured daily cases numbered approximately 1% of the peak. We also checked that in the days following this start date, the daily cases fit well to an exponential function, as would be expected at the start of a pandemic (Appendix A).

The results for r_eff_, f_tot_, L_eff_ and α for six countries which had r_eff_ varying from 1.23 to 6.04 are shown in Figure 2 a-f. The results for the parameters for all thirty-four countries are shown in Table 1. Supplementary Figure 1 shows plot of the data for X(t) and the fits for all thirty-four countries. Also shown in the plots are the location of the start day (caseload = 1% of peak), the location of the peak and of the half width at full maximum as well as the inferred values of L_X_, W_X_ and H_X_ for each country. Some notable exclusions in the list of countries are the United States, the Russian Federation, Canada, India, and Pakistan. The reason for this is that these countries (and others) either had very broad first peaks or had multiple subpeaks within the first peak, making estimates of L_X_ and W_X_ problematic. This is presumably because they cannot be considered homogeneous for a variety of reasons, the most important likely being non-uniform response from authorities regarding the use of masks and variable rules across the country regarding movement of people, quarantine etc. In countries such as the United States and Canada, where the response from the authorities was somewhat state or province specific, it should be possible to do a state-by-state or province-by-province analysis.

**Figure 2.**
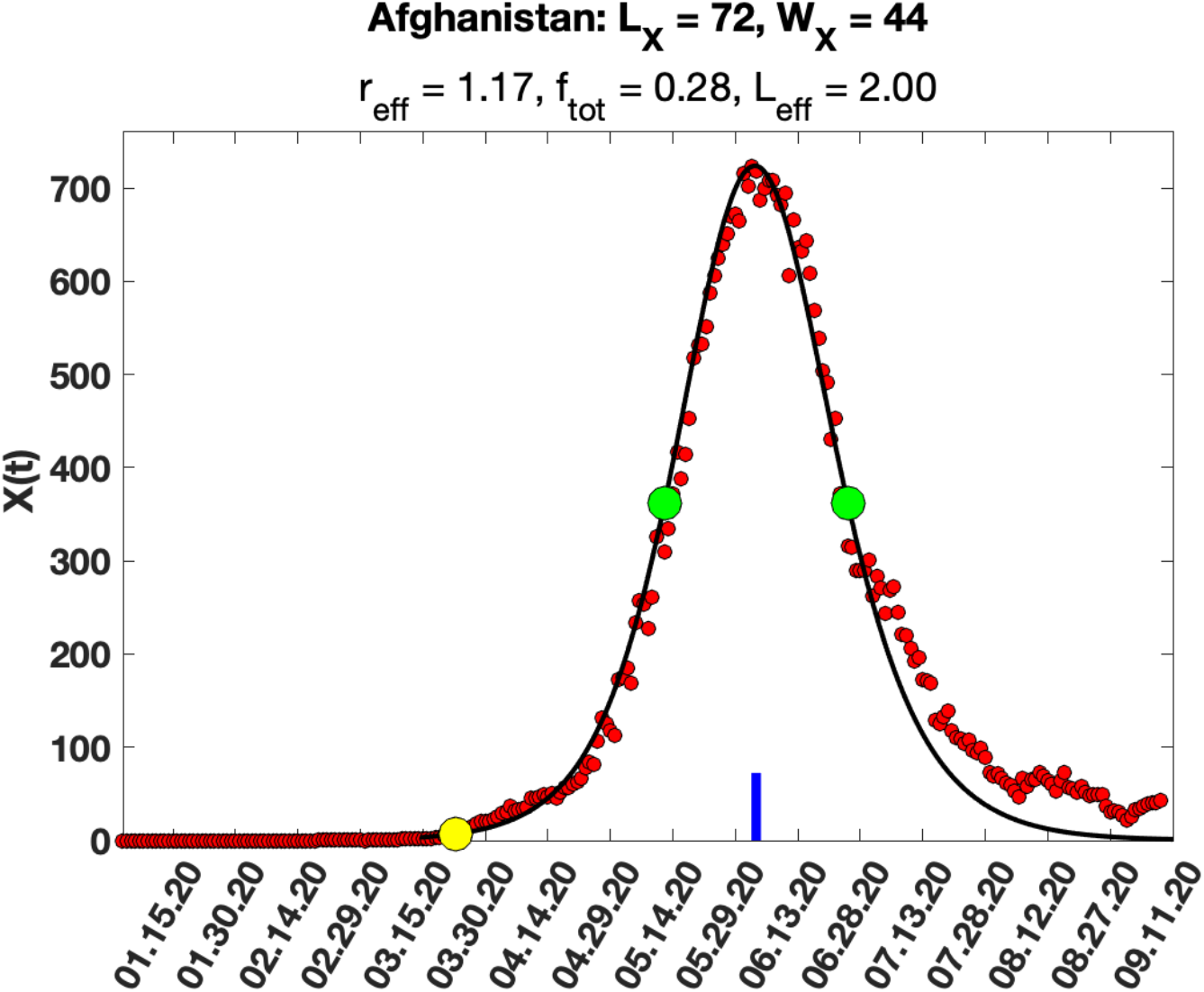
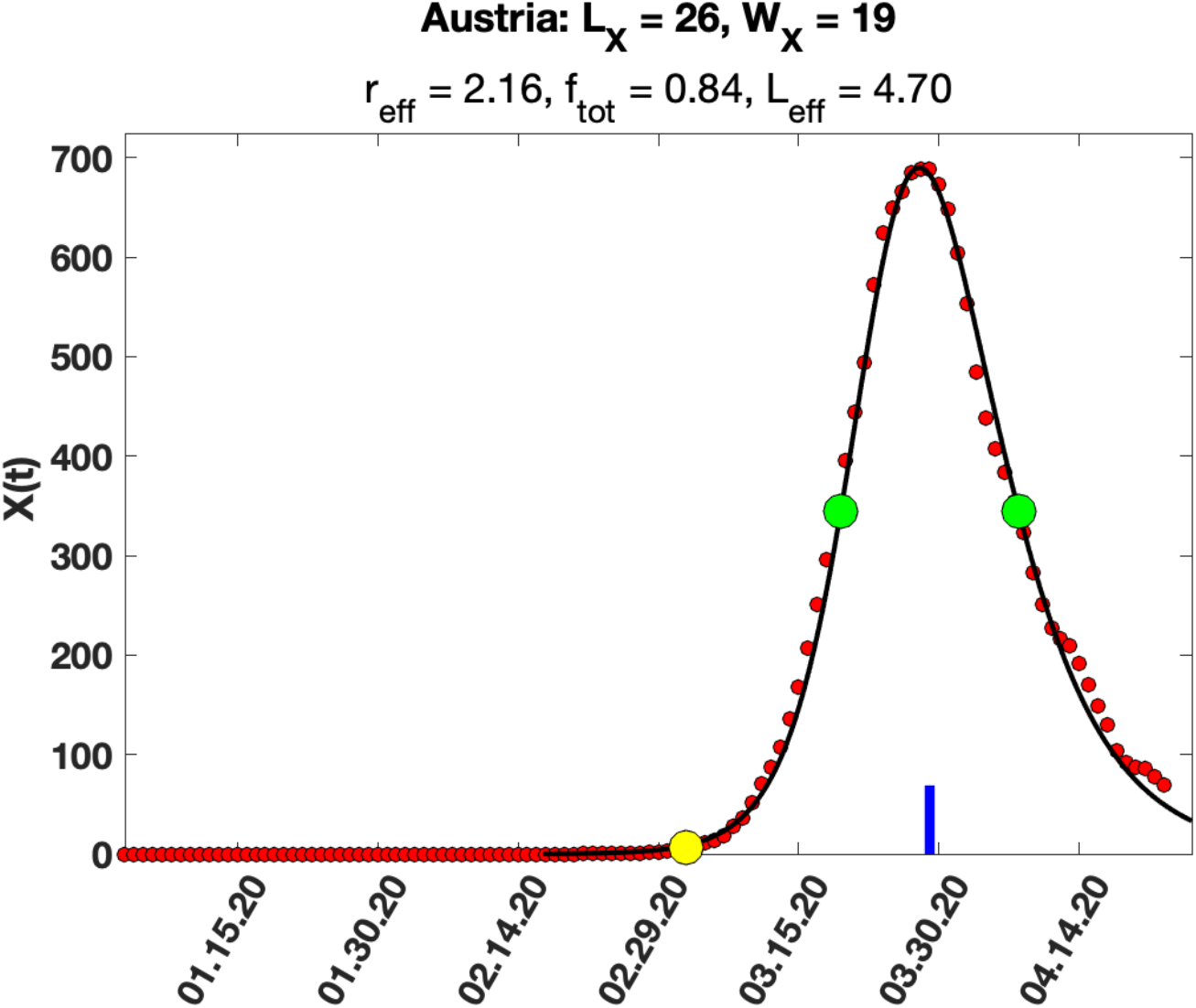
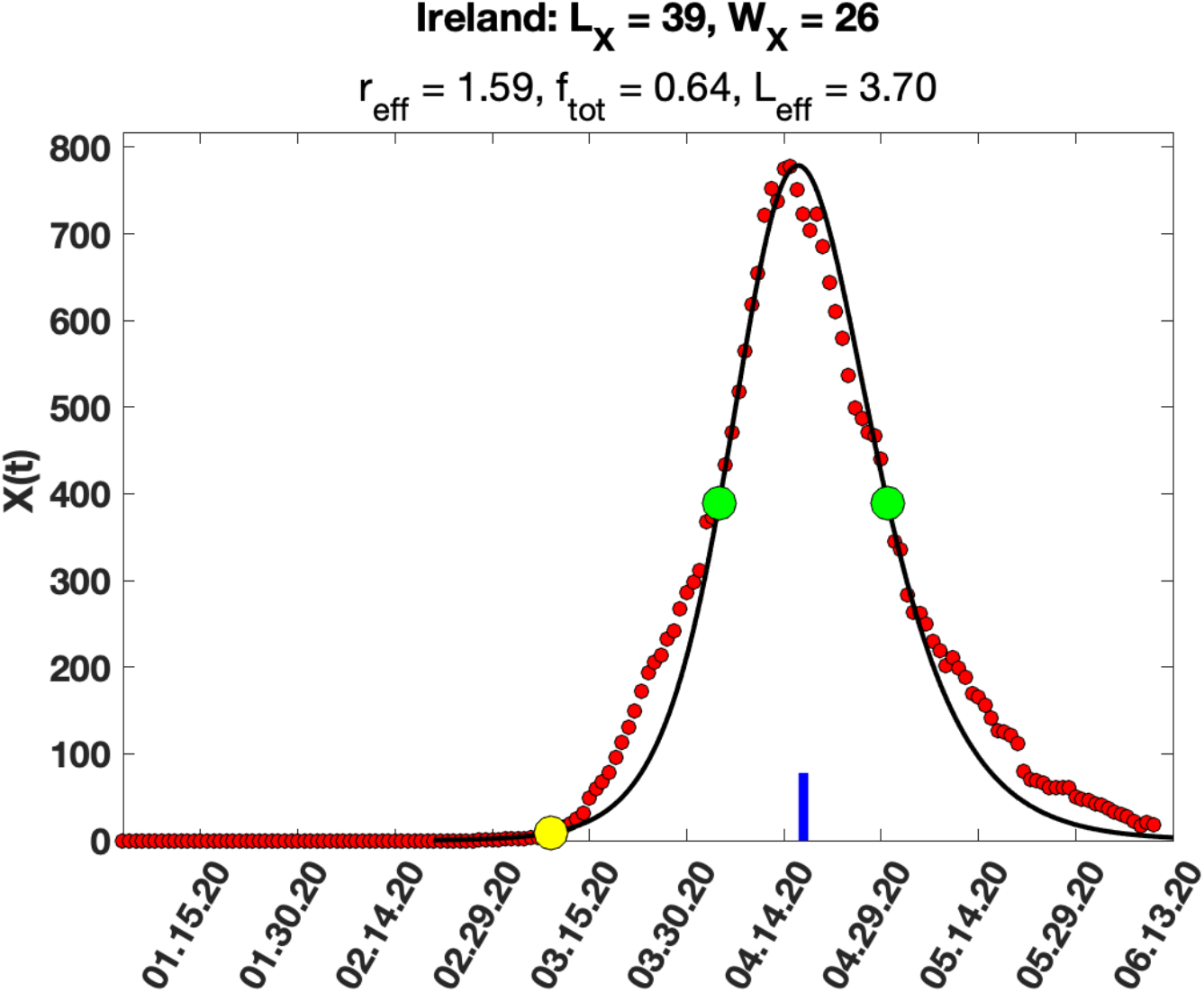
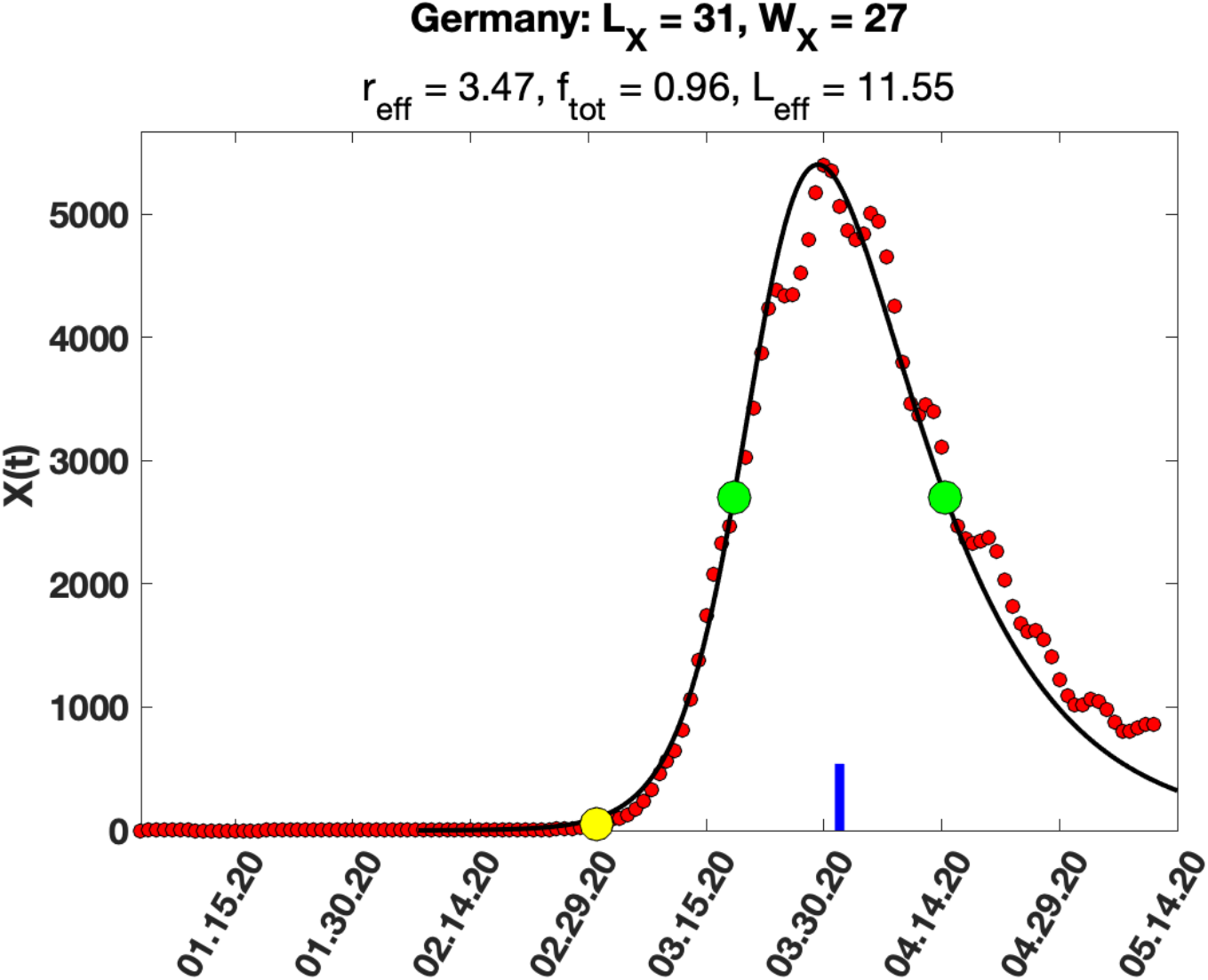
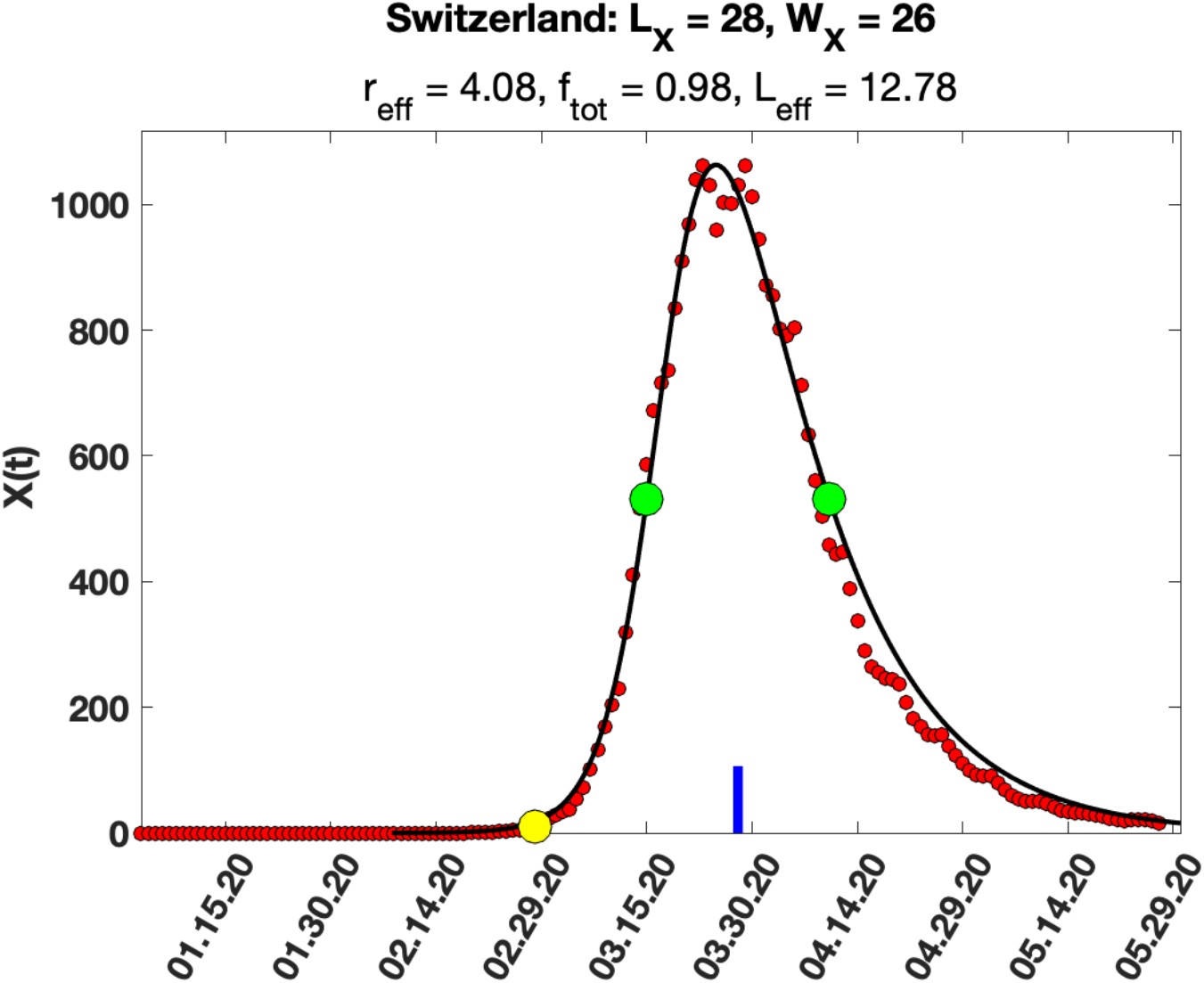
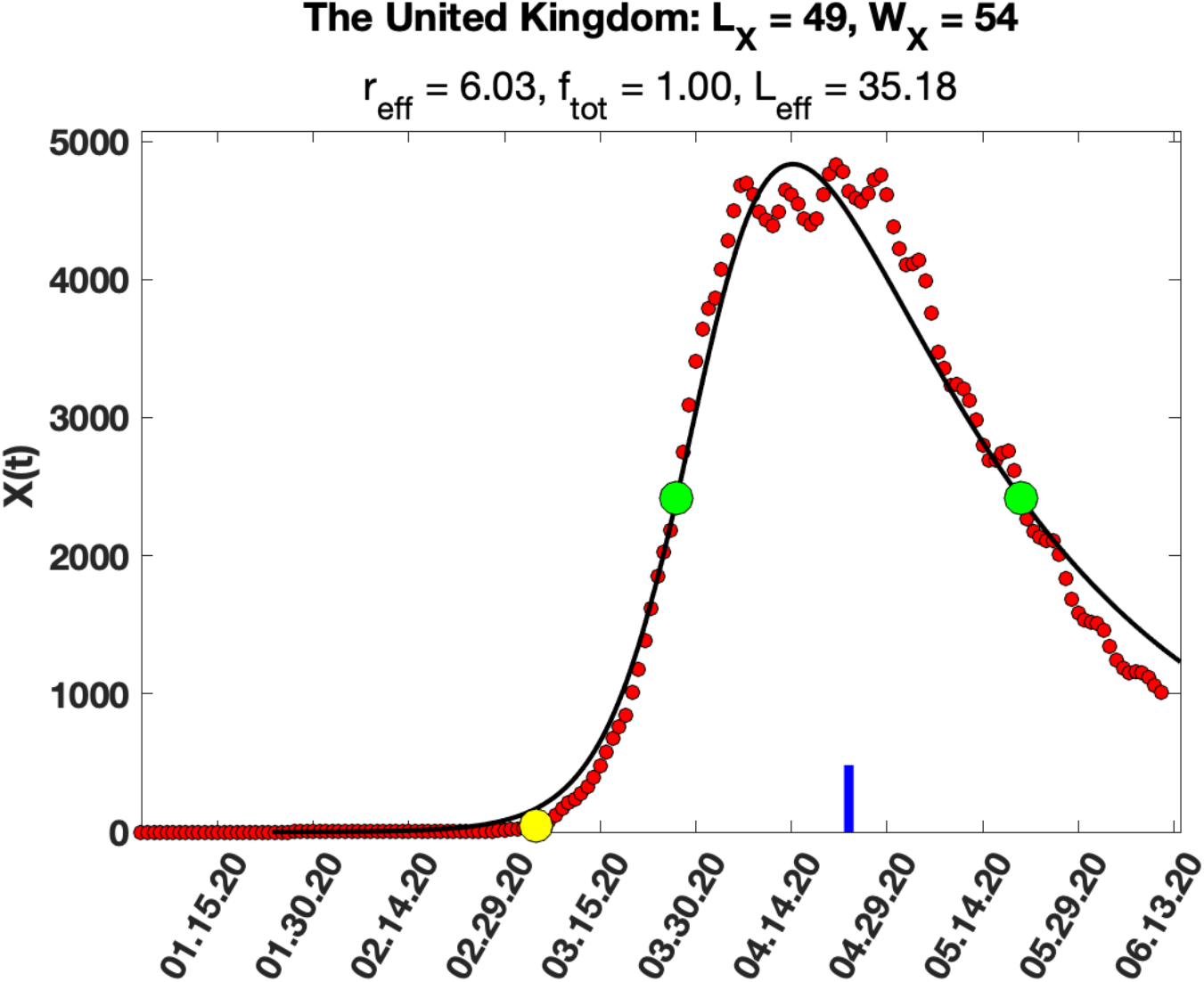
**a-f:** Fits of our model to data for X(t) from the World Health Organization website https://covid19.who.int/data for six of the thirty-four countries for which analysis was possible. The complete set of plots are in Supplementary Figure 1. The red dots are the X(t) data (obtained from the raw data by averaging it over eleven days) and the black curve is the fit obtained at the inferred value of r_eff_, L_eff_ (see text for method). The locations of the “start” of the pandemic (daily cases = 1% of peak) and the location of the maximum in X(t) are shown as a yellow circle and a blue mark on the time axis respectively. The green dots identify the peak half maximum. The values of the fitted parameters are shown in the text above each plot.

**Table 1:** Results for r_eff_, f_tot_, L_eff_ and α (Columns O, Q, S, U) from applying our methods to analyze WHO data (https://covid19.who.int/data) for the first peak in X(t) (daily identified cases) for thirty-four countries which had a clear, well separated peak in X(t) starting January 3, 2020.

| Country | LX<br>(days) | WX<br>(days) | LX/WX | r_eff | f_tot | L_eff<br>(days) | alpha =<br>r_eff/L_eff |
| --- | --- | --- | --- | --- | --- | --- | --- |
| Afghanistan | 72 | 44 | 1.64 | 1.17 | 0.28 | 2.0 | 0.58 |
| Japan | 40 | 25 | 1.60 | 1.27 | 0.39 | 1.8 | 0.72 |
| Australia | 26 | 17 | 1.53 | 1.49 | 0.58 | 2.1 | 0.72 |
| Ireland | 39 | 26 | 1.50 | 1.59 | 0.64 | 3.7 | 0.43 |
| Iceland | 30 | 21 | 1.43 | 1.86 | 0.75 | 4.1 | 0.45 |
| South Africa | 20 | 14 | 1.43 | 1.86 | 0.75 | 2.7 | 0.68 |
| China | 24 | 17 | 1.41 | 1.92 | 0.77 | 3.5 | 0.55 |
| Bolivia | 118 | 85 | 1.39 | 2.06 | 0.81 | 19.6 | 0.10 |
| Austria | 26 | 19 | 1.37 | 2.16 | 0.84 | 4.7 | 0.46 |
| Azerbaijan | 34 | 25 | 1.36 | 2.20 | 0.84 | 6.4 | 0.35 |
| Gambia | 36 | 27 | 1.33 | 2.33 | 0.87 | 7.4 | 0.31 |
| Qatar | 69 | 54 | 1.28 | 2.64 | 0.91 | 17.4 | 0.15 |
| Serbia | 37 | 30 | 1.23 | 2.89 | 0.93 | 10.7 | 0.27 |
| Tajikistan | 27 | 22 | 1.23 | 2.92 | 0.93 | 7.9 | 0.37 |
| France | 35 | 29 | 1.21 | 3.06 | 0.94 | 11.0 | 0.28 |
| Belgium | 38 | 32 | 1.19 | 3.18 | 0.95 | 12.6 | 0.25 |
| Greece | 32 | 27 | 1.19 | 3.19 | 0.95 | 10.6 | 0.30 |
| New Zealand | 20 | 17 | 1.18 | 3.26 | 0.96 | 6.8 | 0.48 |
| Spain | 28 | 24 | 1.17 | 3.35 | 0.96 | 9.9 | 0.34 |
| Czechia | 29 | 25 | 1.16 | 3.40 | 0.96 | 10.5 | 0.32 |
| Germany | 31 | 27 | 1.15 | 3.47 | 0.97 | 11.5 | 0.30 |
| South Korea | 16 | 14 | 1.14 | 3.51 | 0.97 | 6.0 | 0.58 |
| Israel | 31 | 28 | 1.11 | 3.82 | 0.98 | 13.0 | 0.29 |
| Türkiye | 34 | 31 | 1.10 | 3.90 | 0.98 | 14.7 | 0.27 |
| Thailand | 23 | 21 | 1.10 | 3.91 | 0.98 | 10.0 | 0.39 |
| Cuba | 37 | 34 | 1.09 | 3.97 | 0.98 | 16.4 | 0.24 |
| Switzerland | 28 | 26 | 1.08 | 4.08 | 0.98 | 12.8 | 0.32 |
| Norway | 27 | 26 | 1.04 | 4.49 | 0.99 | 13.8 | 0.33 |
| Croatia | 30 | 29 | 1.03 | 4.52 | 0.99 | 15.5 | 0.29 |
| Italy | 40 | 40 | 1.00 | 4.88 | 0.99 | 22.6 | 0.22 |
| Malaysia | 34 | 34 | 1.00 | 4.88 | 0.99 | 19.2 | 0.25 |
| Netherlands | 37 | 37 | 1.00 | 4.88 | 0.99 | 20.9 | 0.23 |
| The United Kingdom | 49 | 54 | 0.91 | 6.03 | 1.00 | 35.2 | 0.17 |
| Portugal | 33 | 37 | 0.89 | 6.25 | 1.00 | 24.6 | 0.25 |

## DISCUSSION

In this paper we have developed a method, applicable to any pandemic, to identify the fraction of infected individuals from among the pool of interacting susceptible individuals in a given region, using a simple extension of the epidemiological SIR model [30]. We show that in this model, there is a universal scaling function that relates the ratio of the location L_X_, and the width W_X_ of the peak in daily identified cases X(t) to the effective Pandemic R-parameter r_eff_ and the fraction f_tot_ of infected exposed individuals (including both symptomatic and asymptomatic infected individuals) (see Figure 1 and Supplementary Table 1). This in turn allows an estimate of the effective latency L_eff_ (average number of days an infected individual is able to infect others) and the infection probability α of transmission from an infected individual to a susceptible individual in a single encounter (see Appendix B for details). Within the limits of the SIR model, our results are general and apply to any pandemic. We apply our method to worldwide country specific data to find r_eff_, f_tot_, L_eff_ and α for the first phase (first peak in daily cases) for the SARS-COV-2 pandemic for thirty-four countries which had a clear, well separated peak in daily cases (Table 1, Figure 2, Supplementary Figure 2).

*It is important to note that our result for f*_*tot*_ *represents only the fraction of infected individuals in the “exposed population” in a given region – i*.*e*., *it only applies to the set of susceptible individuals who came into sufficiently close contact with infected individuals for the virus to transmit. This value should not be taken to represent the fraction of infected individuals in the population as a whole, because our analysis does not include those individuals who were sufficiently isolated in some way (e*.*g*., *self-quarantined, wore masks etc*.*), so as to avoid any contact with the virus*.

With this caveat in mind, we note that our results suggest that in the SARS-COV-2 pandemic, the fraction of infected individuals who were exposed to the virus was very high in most countries that met our analysis criteria, suggesting that in its early stages, when countries did not impose quarantines and the use of masks was limited, this virus was highly effective in transmission. In some of the developed countries, our results suggest that almost all exposed individuals were infected in the first phase of the pandemic (Table 1). The only countries where f_tot_ was less than 0.5 were those with a low population density (Australia), low mobility rates of citizens (Afghanistan) or where the use of masks was common, even in the absence of a pandemic (Japan).

Several countries, notably the United States, Canada, The Russian Federation, India, and Pakistan did not meet our criterion of a clear, well separated first peak in daily cases in 2020. This is most likely due to the fact that they cannot be thought of as homogeneous in the sense of response from local authorities regarding the use of masks, quarantine etc. In the United States for example, the response was state and/or county specific. In principle, our method could be applied at the county or state level in the US or the province or sub-province level in Canada to determine parameters from recorded case data, if these compartments had uniform rules for containment of the virus.

Our method can also be applied to subsequent recurrences of the SARS-COV-2 virus (second, third, fourth peaks in daily cases), as the virus evolved into less virulent and more infective strains. Comparing changes in the inferred parameters across countries would provide a country specific overall estimate of preventive measures, such as the effectiveness/efficacy of vaccination, changes in behavior (mask use, testing/quarantine, work-from-home, social distancing, travel restrictions) etc. Furthermore, this method can be applied to other viral pandemics, such as the SARS-COV pandemic of 2003, and Influenza pandemics of the past, such as the H1N1 Spanish Flu pandemic of 1918-19 which recurred in 1950 and 1977, the H2N2 Asian Flu pandemic of 1957, the H3N2 Hongkong pandemic of 1968 and the more deadly H5N1 East Asian pandemic of 1997.

## Data Availability

The data and Matlab codes used in this paper are available on request to. The data used to fit the model to actual pandemic data is in Supplementary Table 1. The World Health Organization country specific data we used for the SARS-CoV-2 pandemic is in Supplementary Table 2. The data in Supplementary Figures and Tables is available online at: https://drive.google.com/drive/folders/1Xai4qk0n4BN7yJvwWBrtssv5YjfyBBTj?usp=sharing

https://drive.google.com/file/d/1P6emrvTBMC0uo-dD6I21U2_iv1tjK2hS/view?usp=sharing

## Data Availability

The data for the universal scaling laws relating the location and width of the peak in daily recorded cases to pandemic parameters r_eff_ and f_tot_, obtained by numerical solution of Eq 6-10 is in Supplementary Table 1. The World Health Organization country specific data the SARS-CoV-2 pandemic which was used to demonstrate the utility of the method is in Supplementary Table 2.

### Author Approval

All authors have seen and approved the manuscript.

## Acknowledgements

GB was partly supported by grants from DoD (KC180159) and NIH (P01CA250957). GB thanks Professor Charles DeLisi for many discussions and collaboration on earlier (unpublished) work on the SARS-CoV-2 pandemic using the SIR model. GB and SD thank the Aspen Center for Physics, which is supported by National Science Foundation grant PHY-1607611, and the Geballe Lab for Advanced Materials at Stanford University for their hospitality while this work was done.

## Supplementary Figure and Table Captions

**Supplementary Figure 1:** Fits of our model to data for X(t) from the World Health Organization website https://covid19.who.int/data for thirty-four countries for which analysis was possible. The red dots are the X(t) data (obtained from the raw data by averaging it over eleven days) and the black curve is the fit obtained at the inferred value of r_eff_, L_eff_ (see text for method). The locations of the “start” of the pandemic (daily cases = 1% of peak) and the location of the maximum in X(t) are shown as a yellow circle and a blue mark on the time axis respectively. The green dots identify the peak half maximum. The values of the fitted parameters are shown in the text above each plot.

**Supplementary Table 1:** Results obtained by numerically solving Eq. 6-8 using the stiff ODE solver ode15s in Matlab for r_eff_ in the range 0.5-6.5. These data were used to derive all the results in the paper.

**Supplementary Table 2:** World Health Organization data for the SARS-CoV-2 pandemic from https://covid19.who.int/data that was used in our analysis.

