## Supplementary figures and images for "Scaling rules for pandemics: Estimating infected fraction from identified cases for the SARS-Cov-2 Pandemic"

### Afghanistan.png

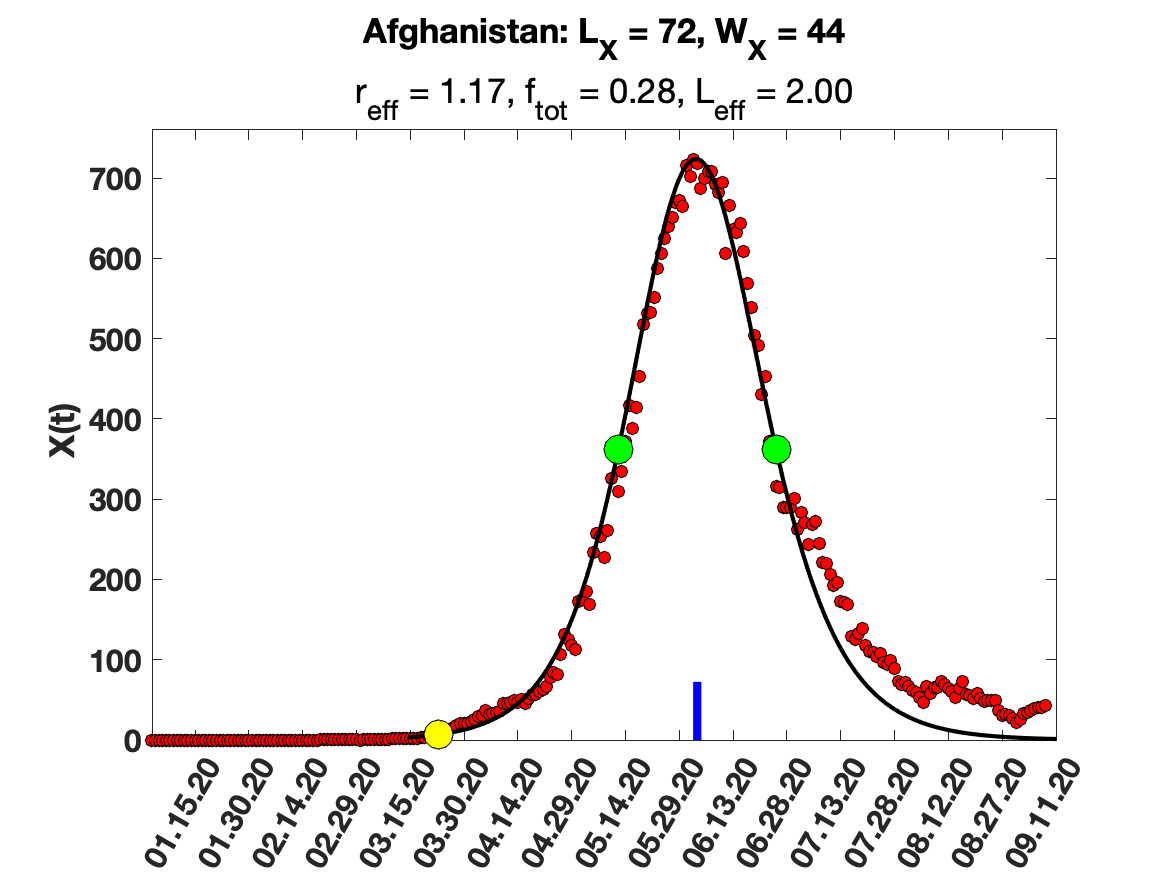

### Australia.png

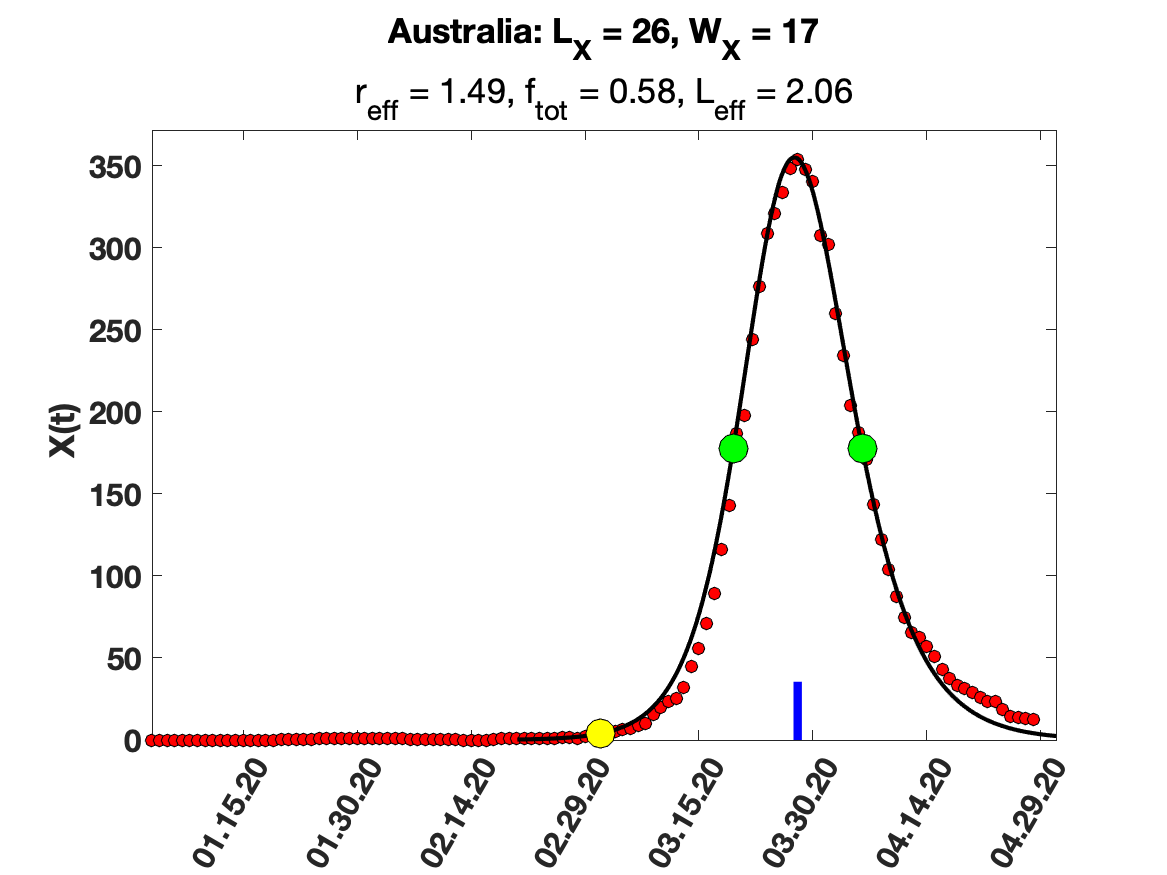

### Austria.png

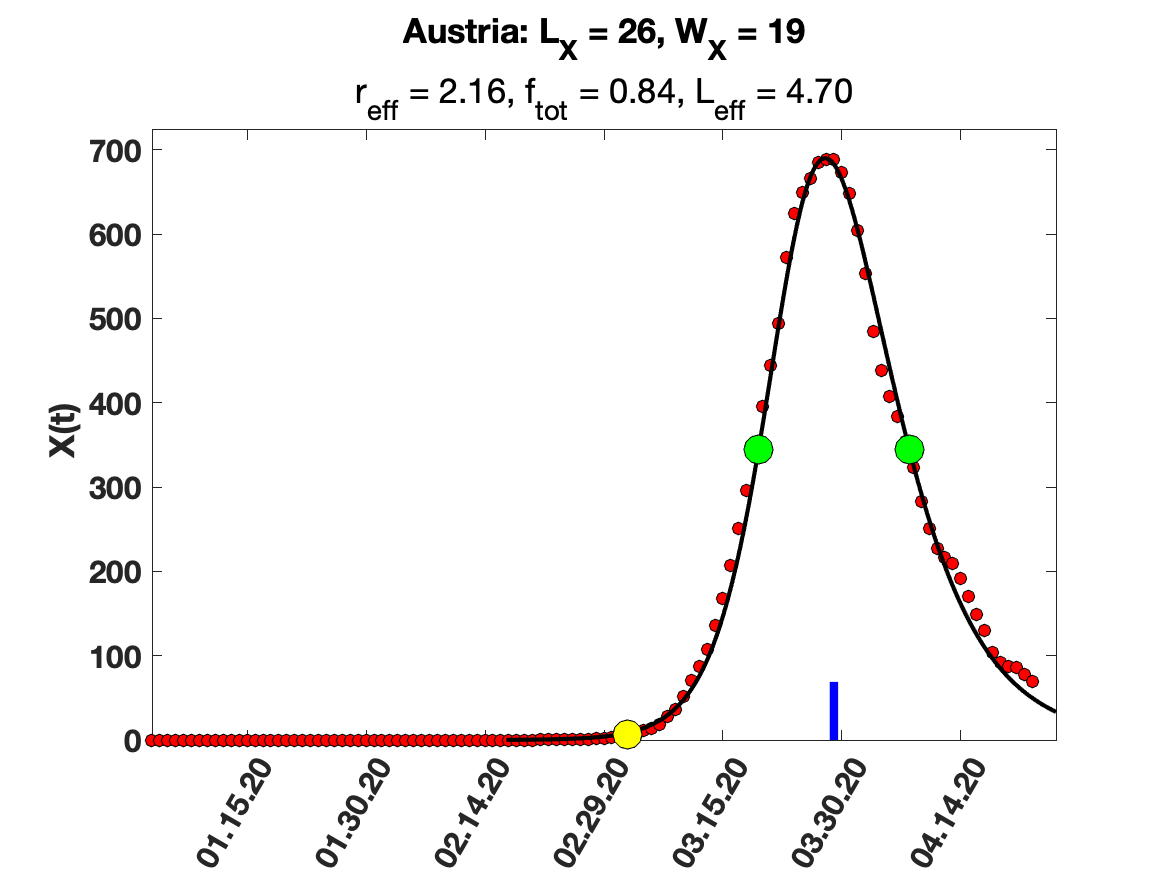

### Azerbaijan.png

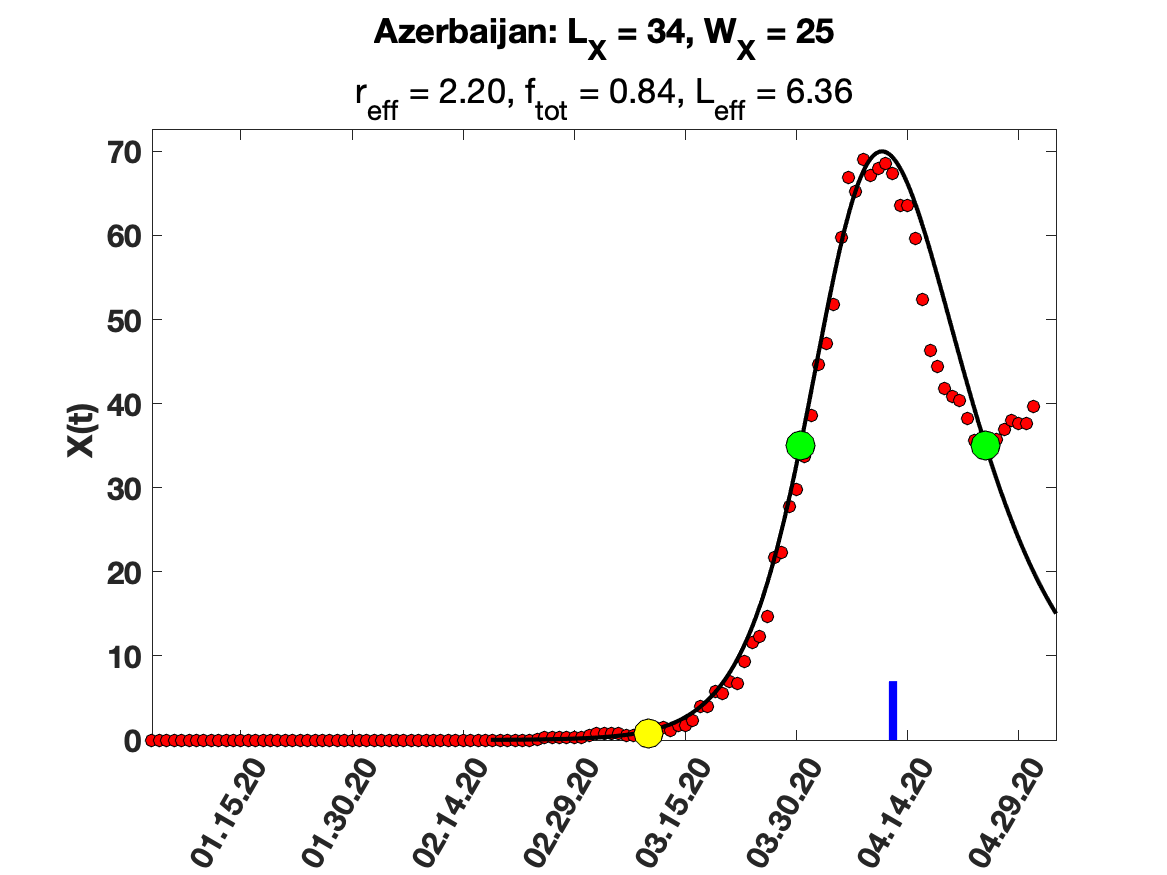

### Belgium.png

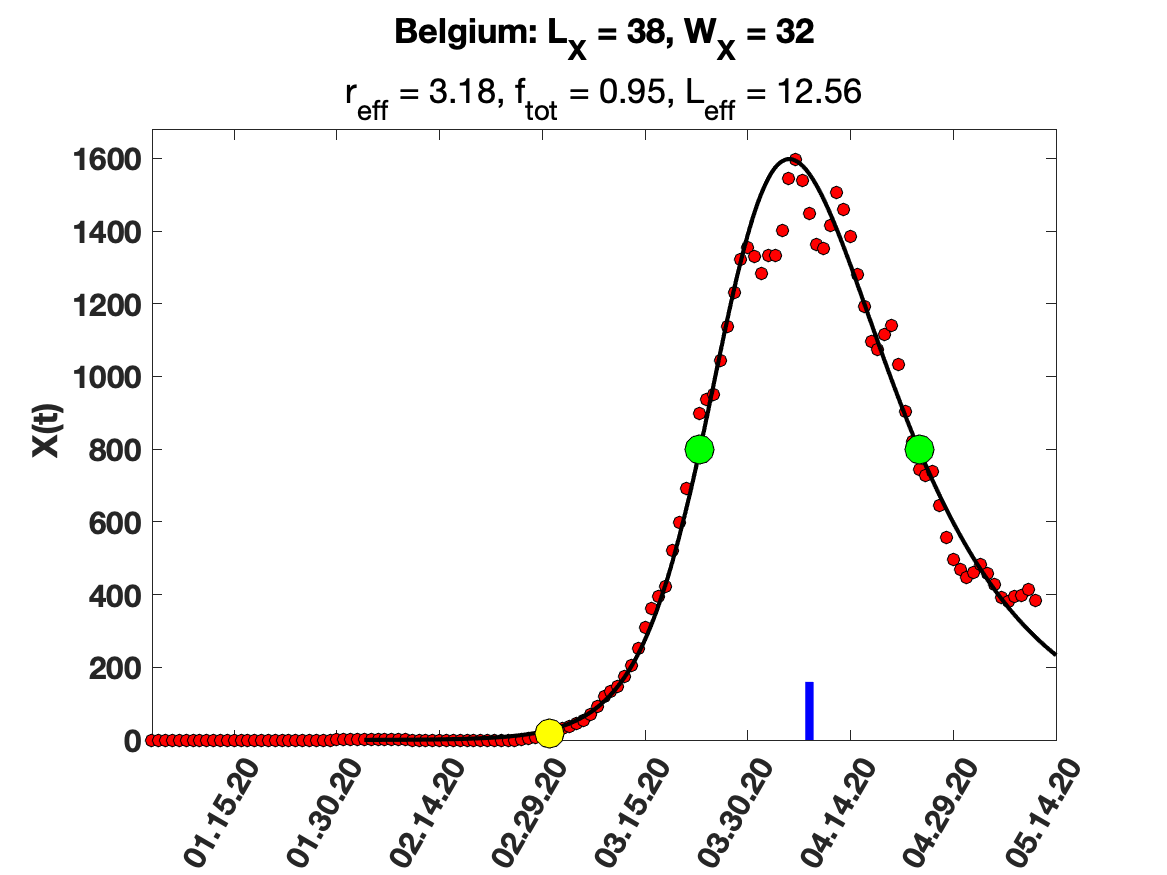

### Bolivia.png

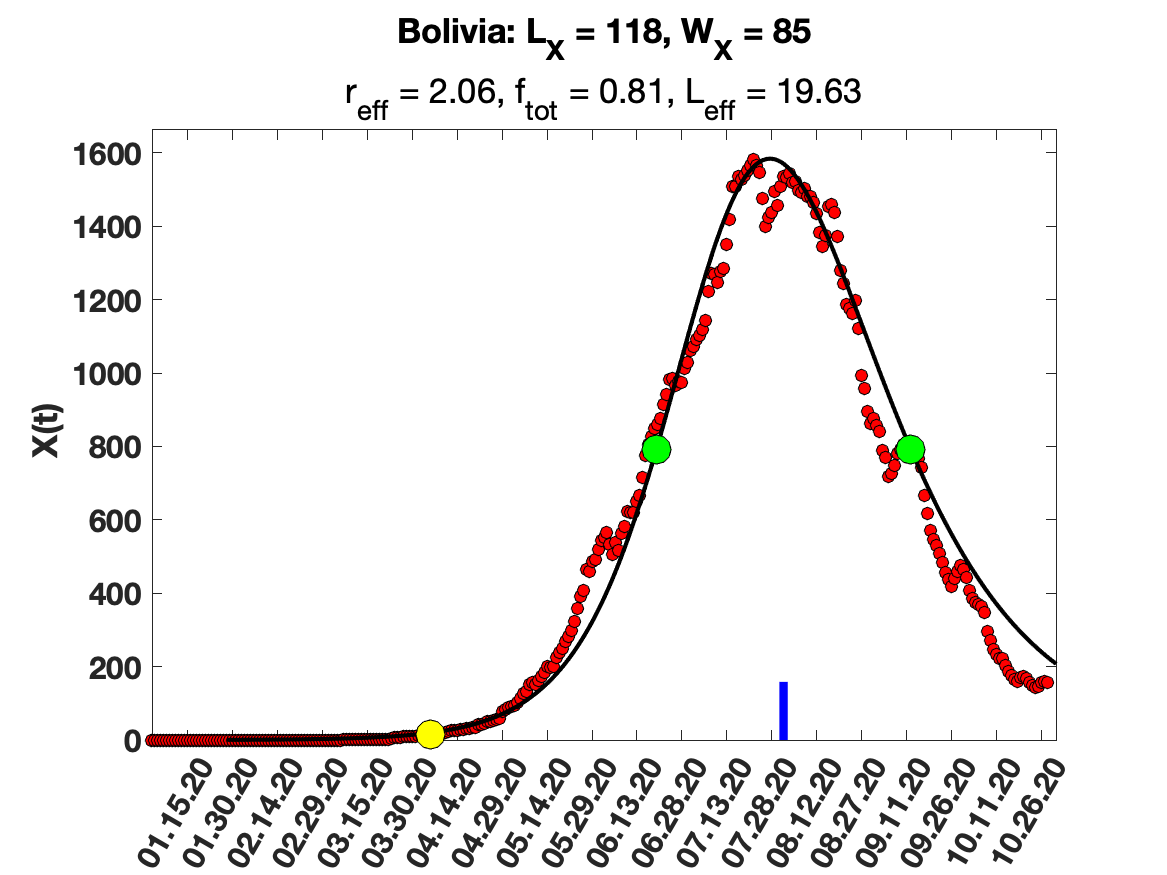

### China.png

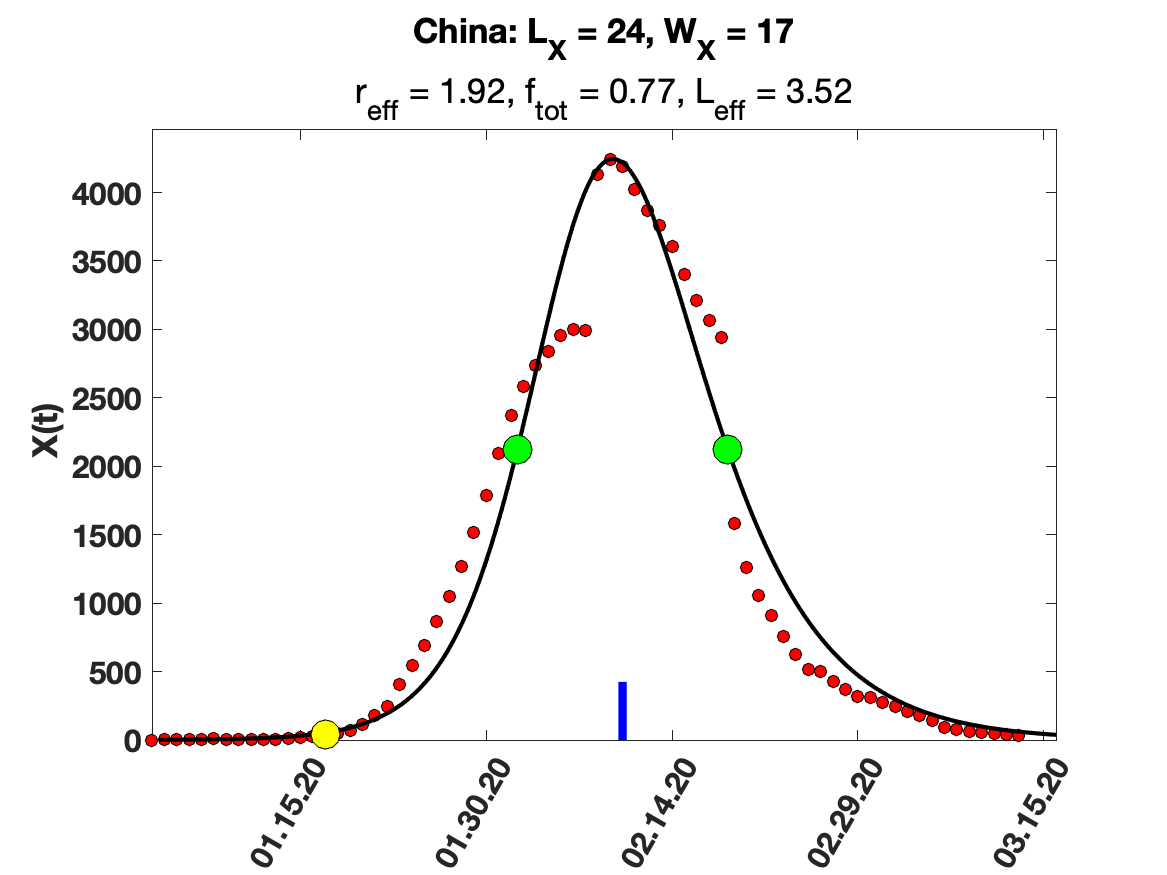

### Croatia.png

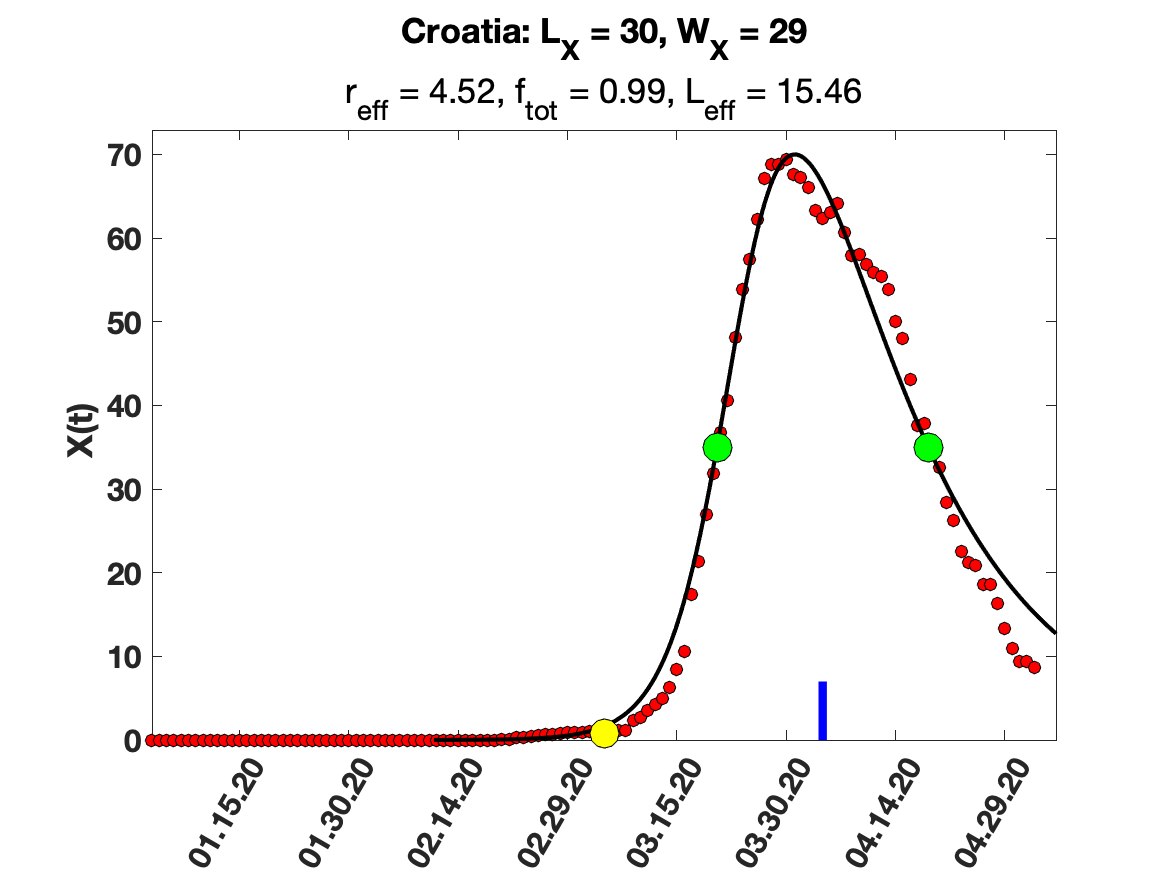

### Cuba.png

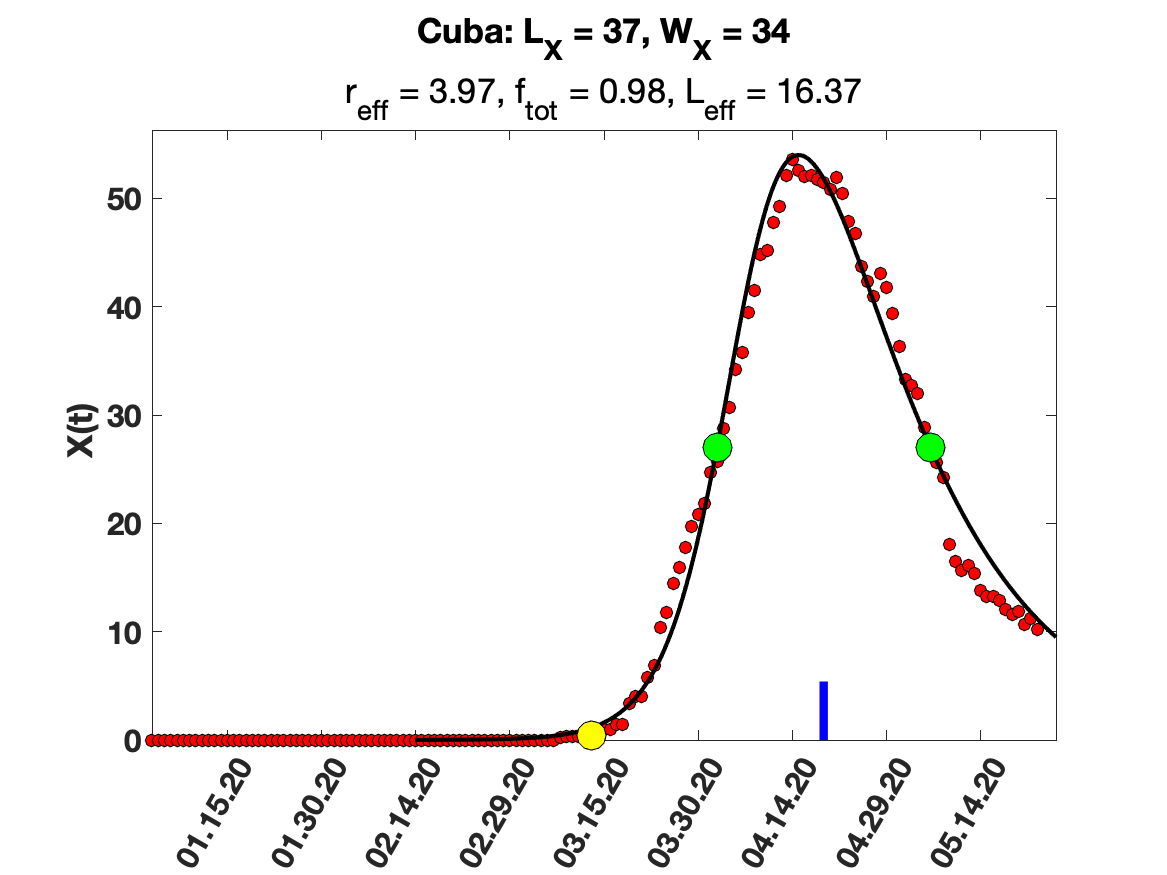

### Czechia.png

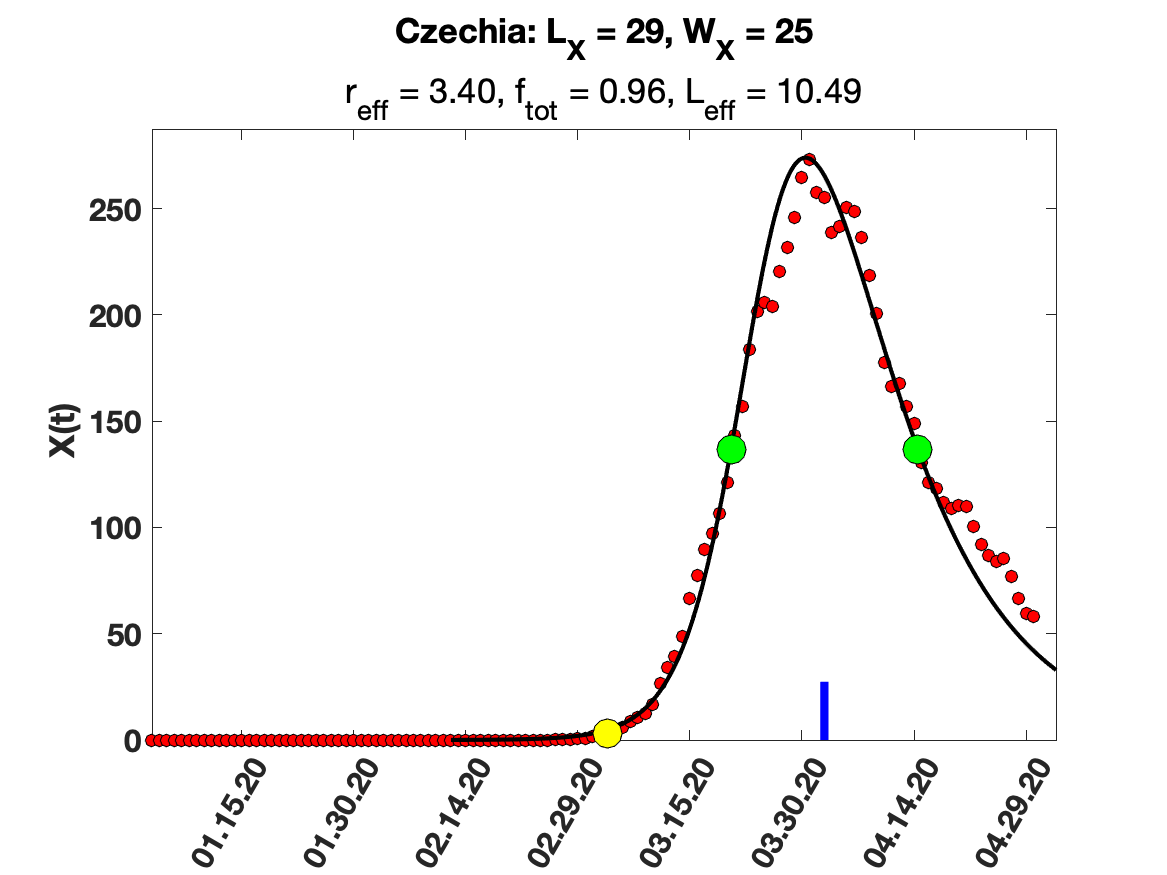

### France.png

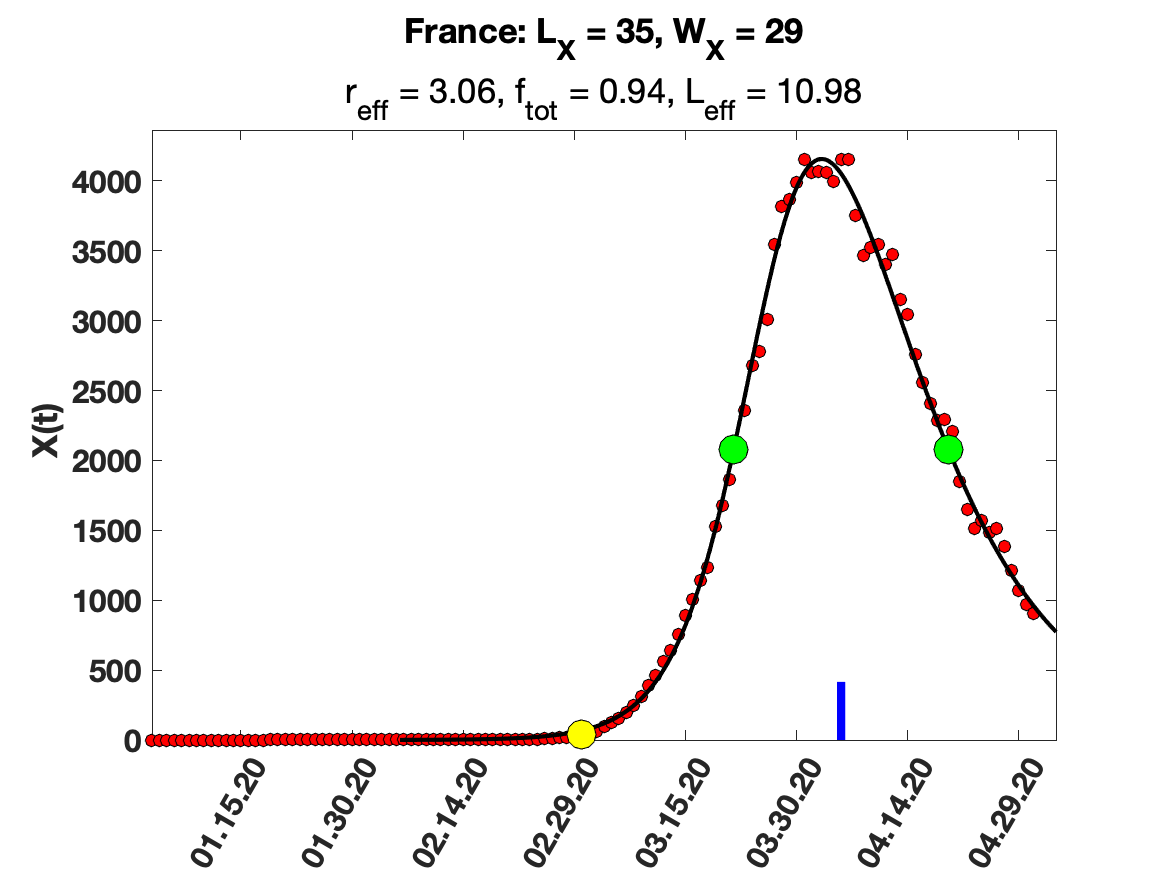

### Gambia.png

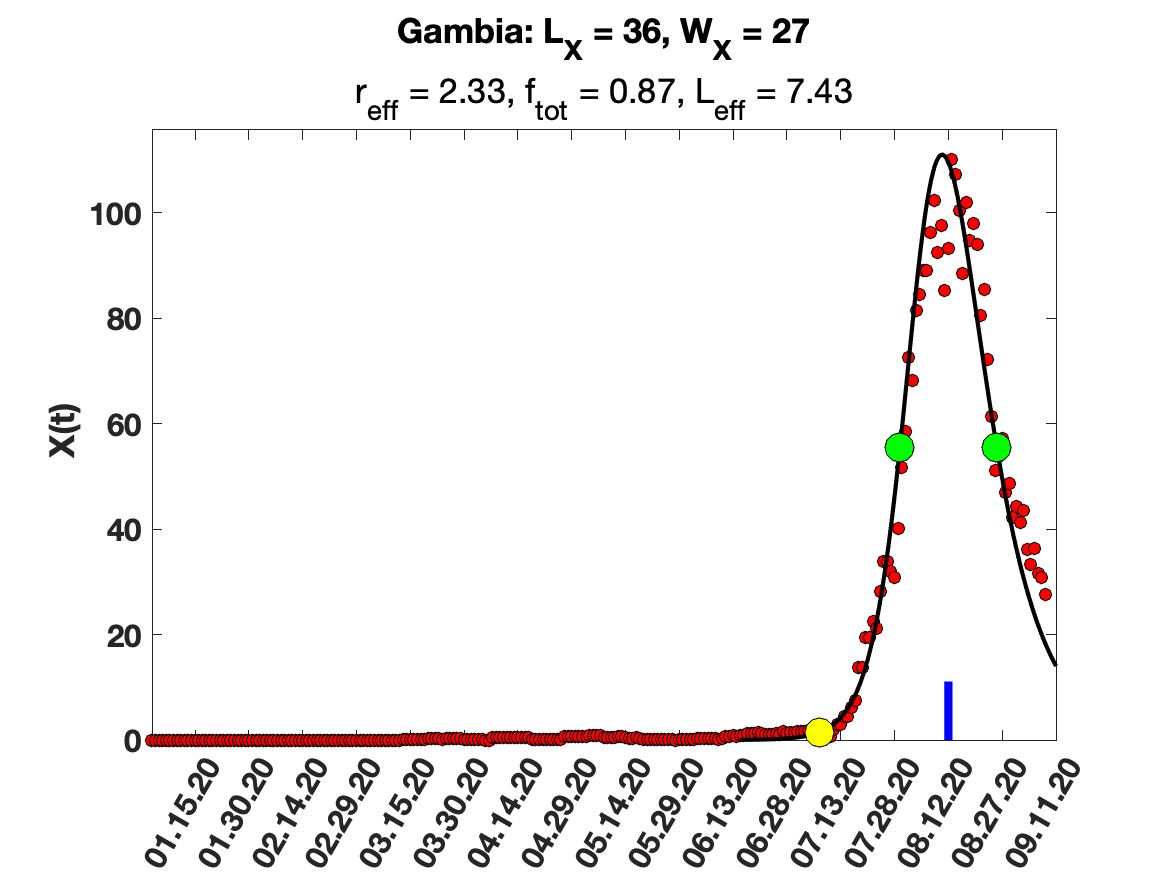

### Germany.png

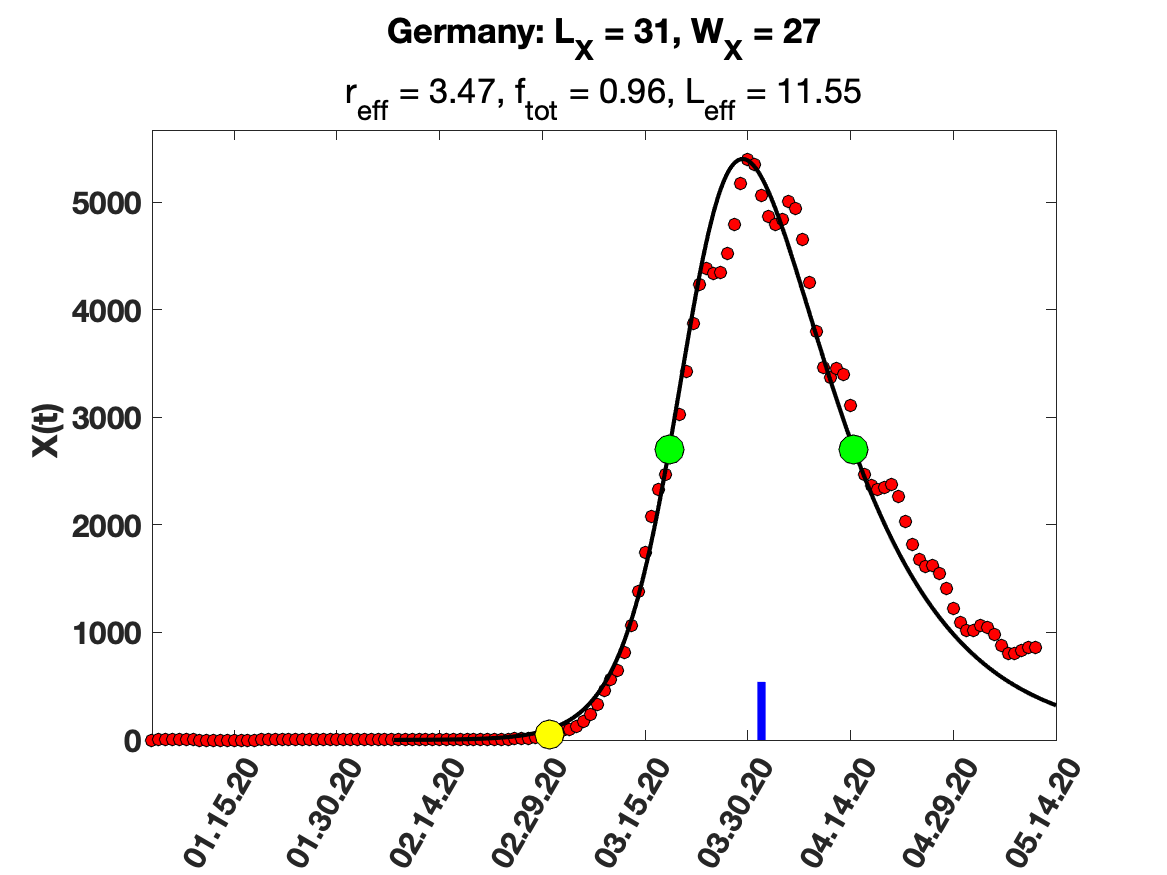

### Greece.png

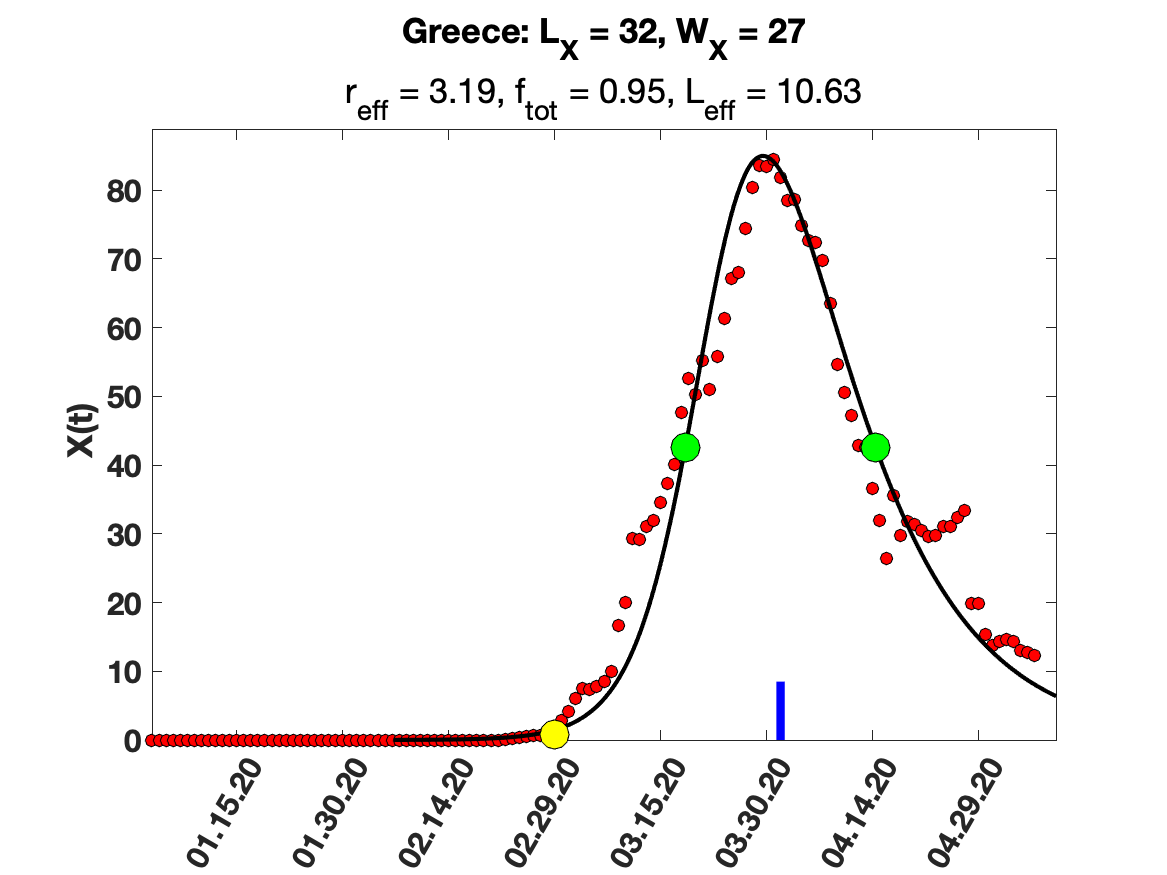

### Iceland.png

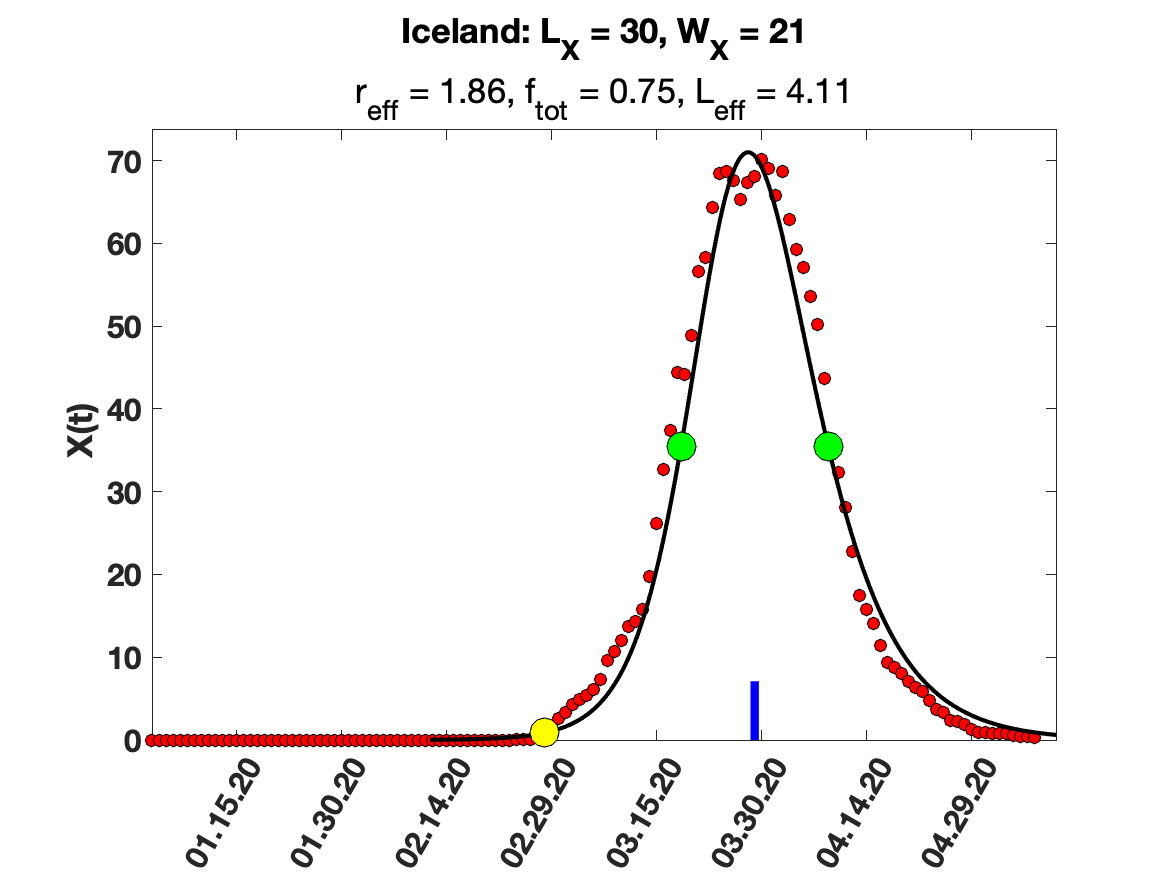

### Ireland.png

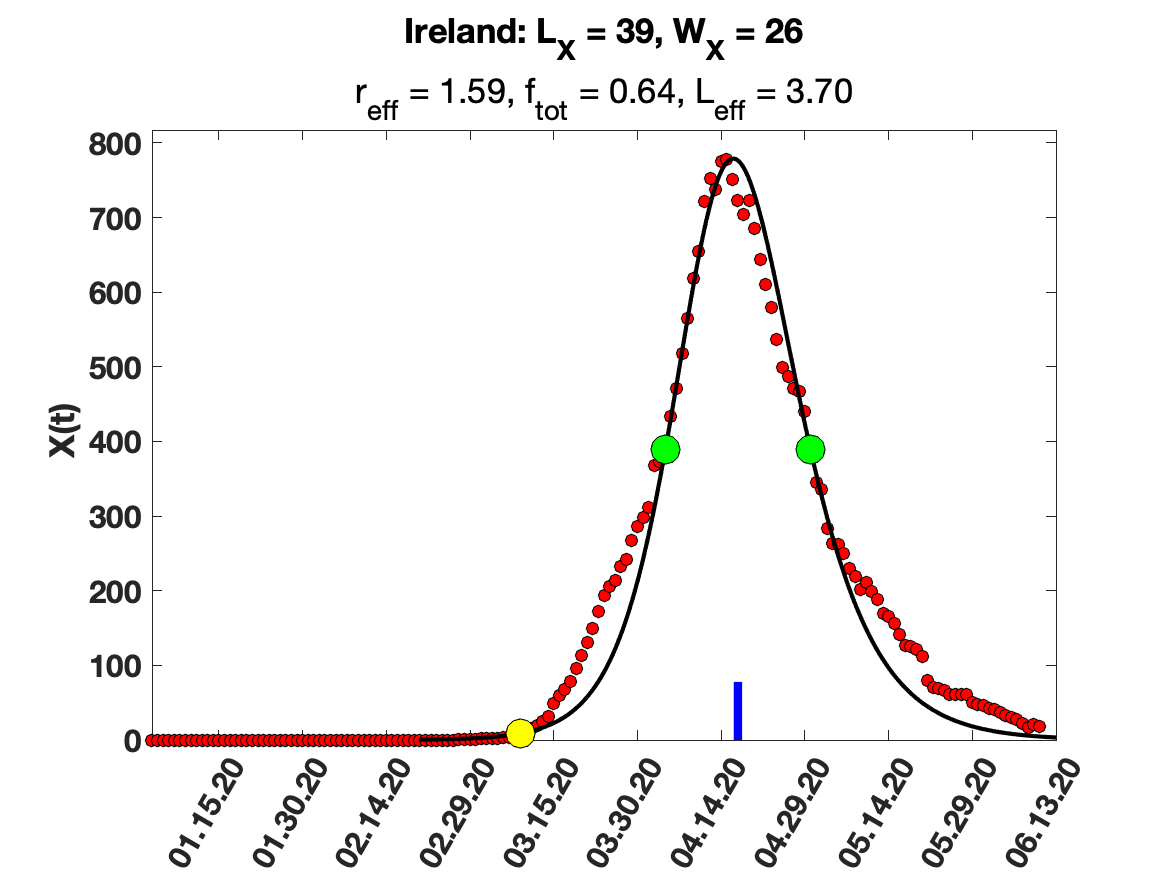

### Israel.png

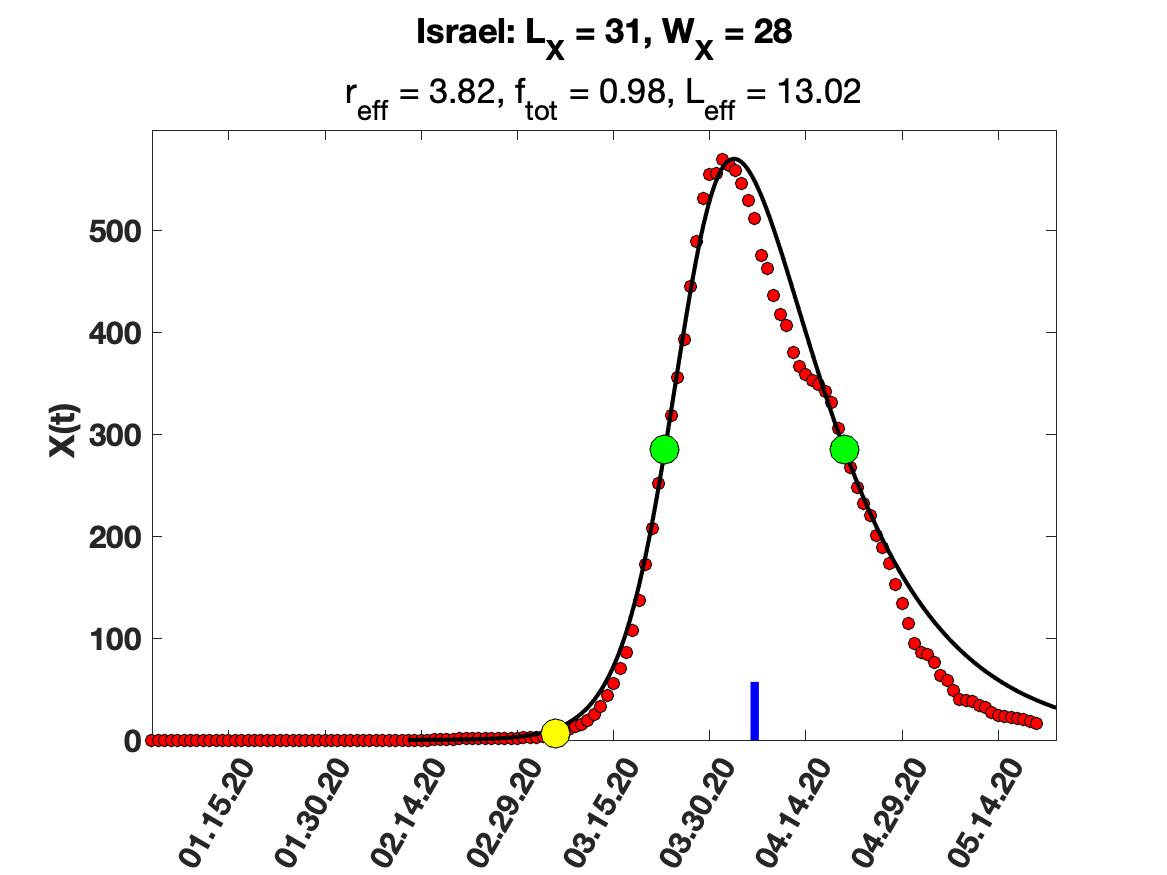

### Italy.png

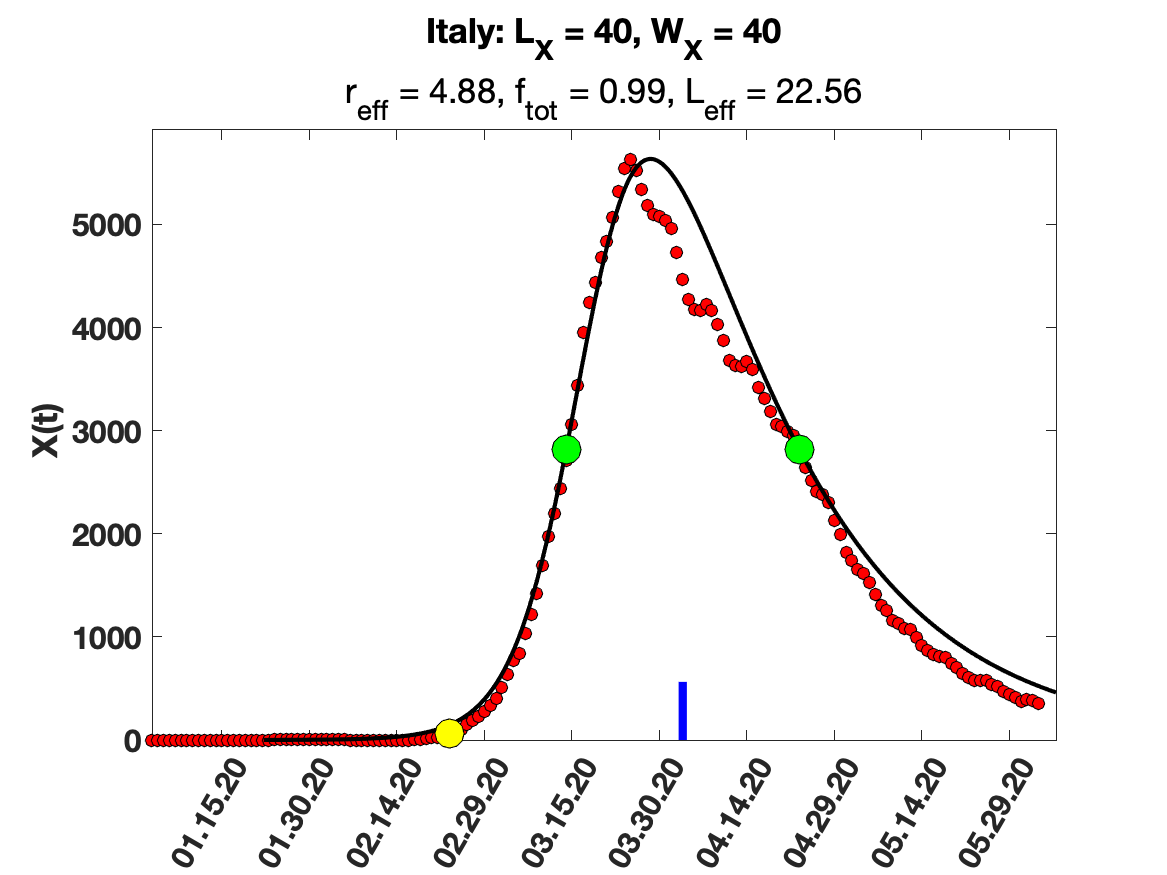

### Japan.png

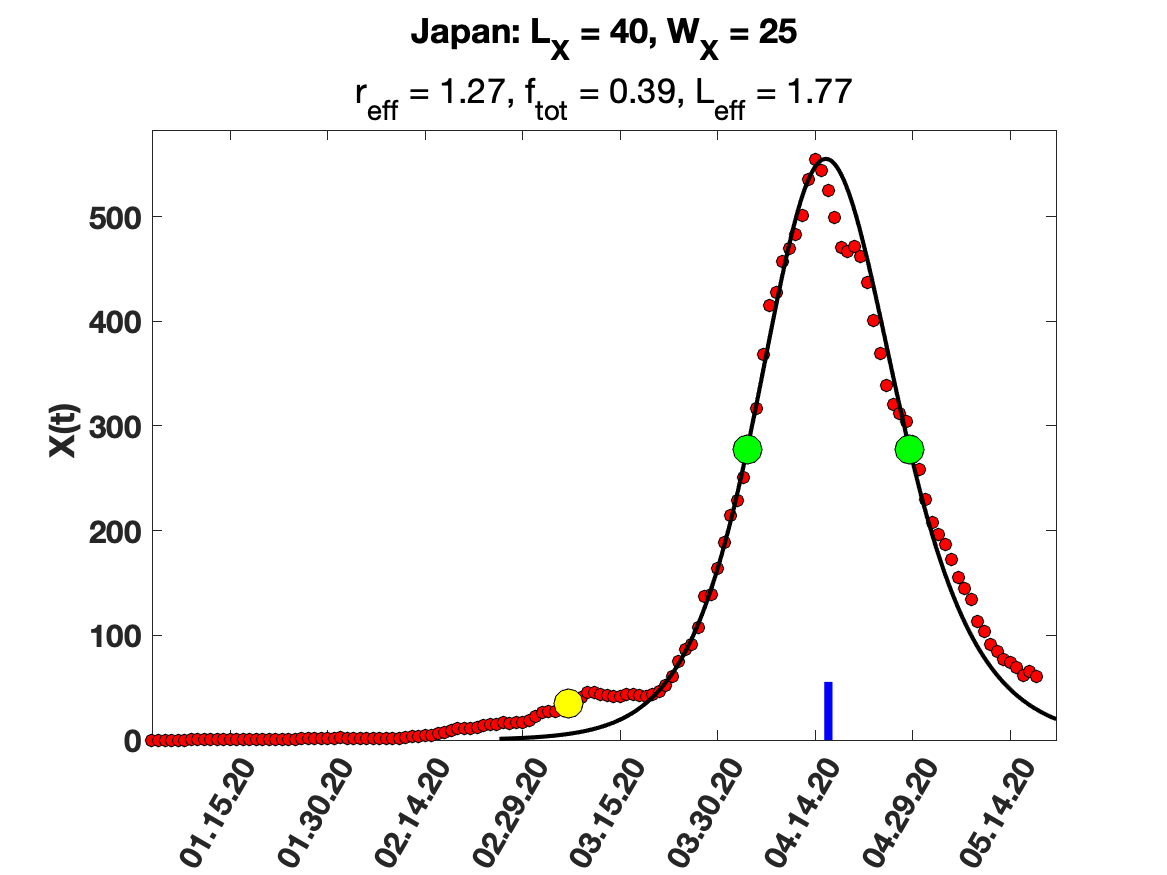

### Malaysia.png

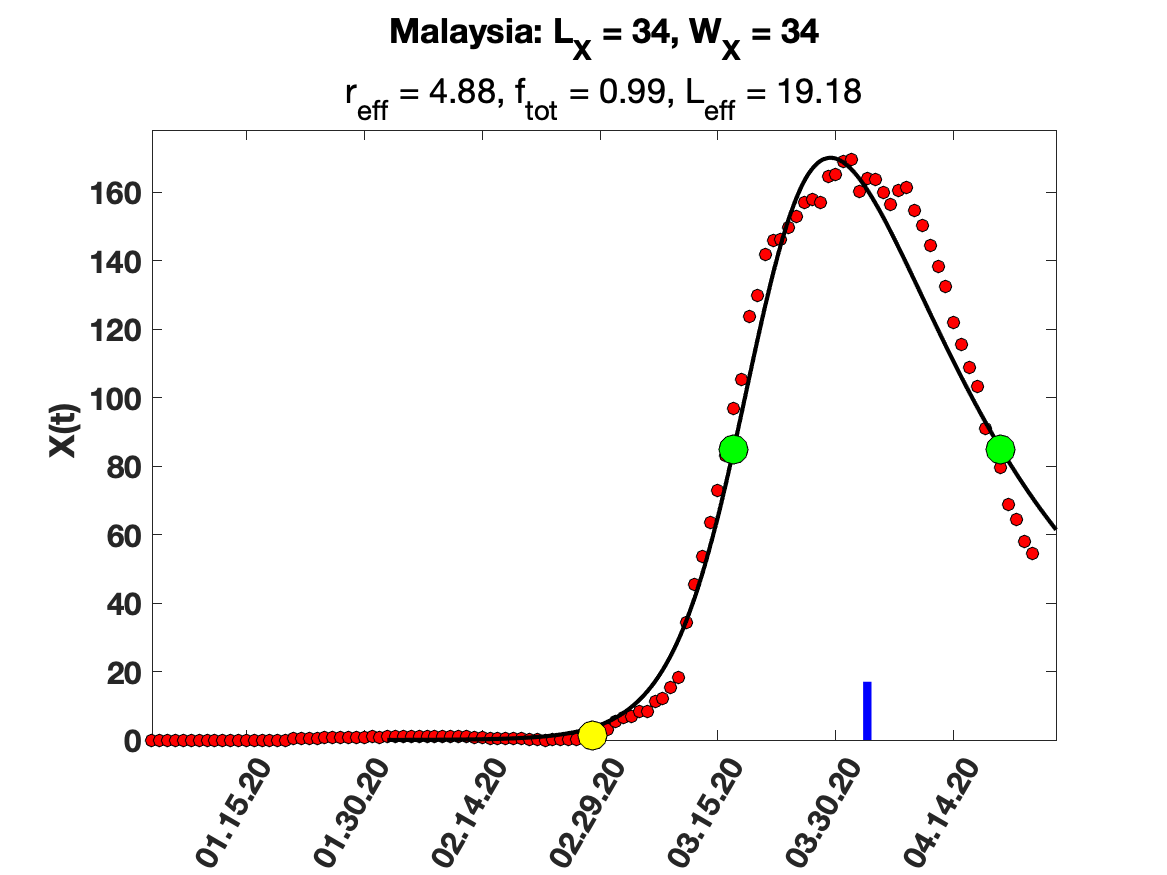

### Netherlands.png

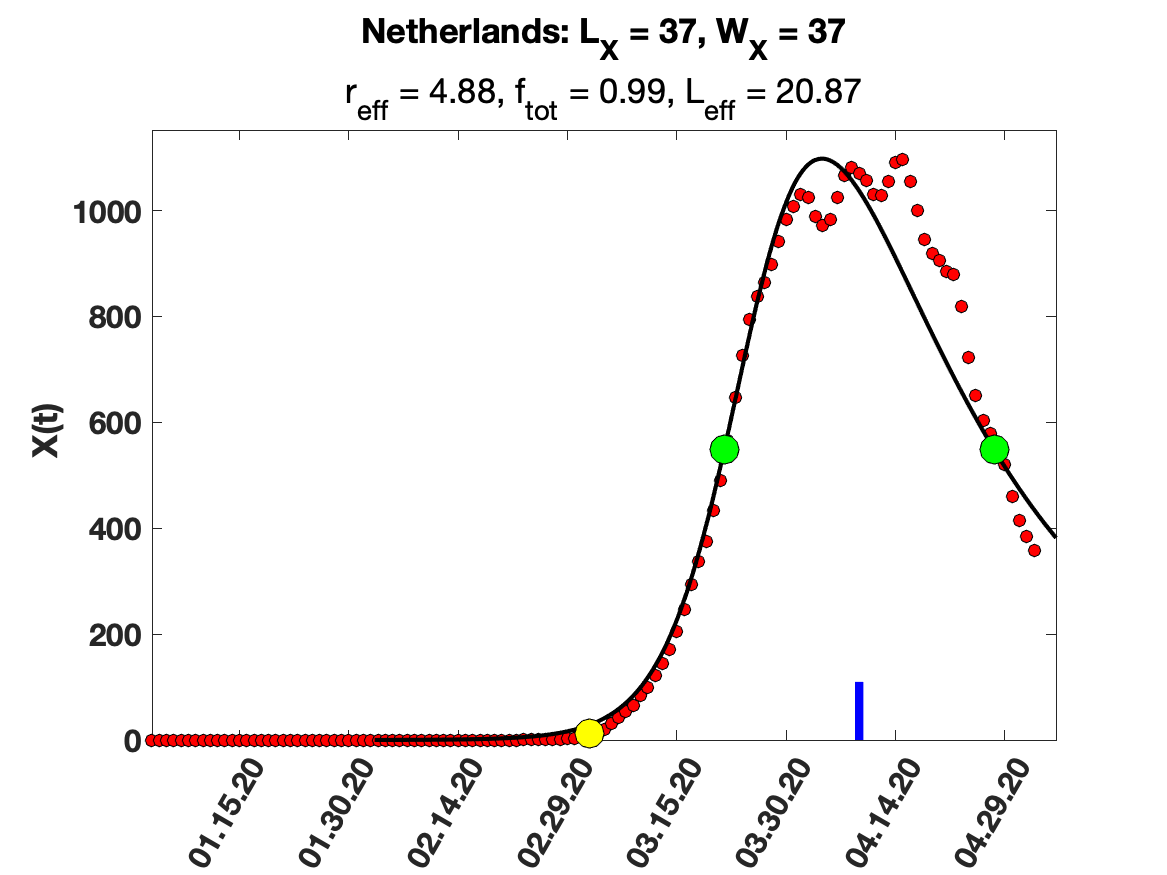

### New Zealand.png

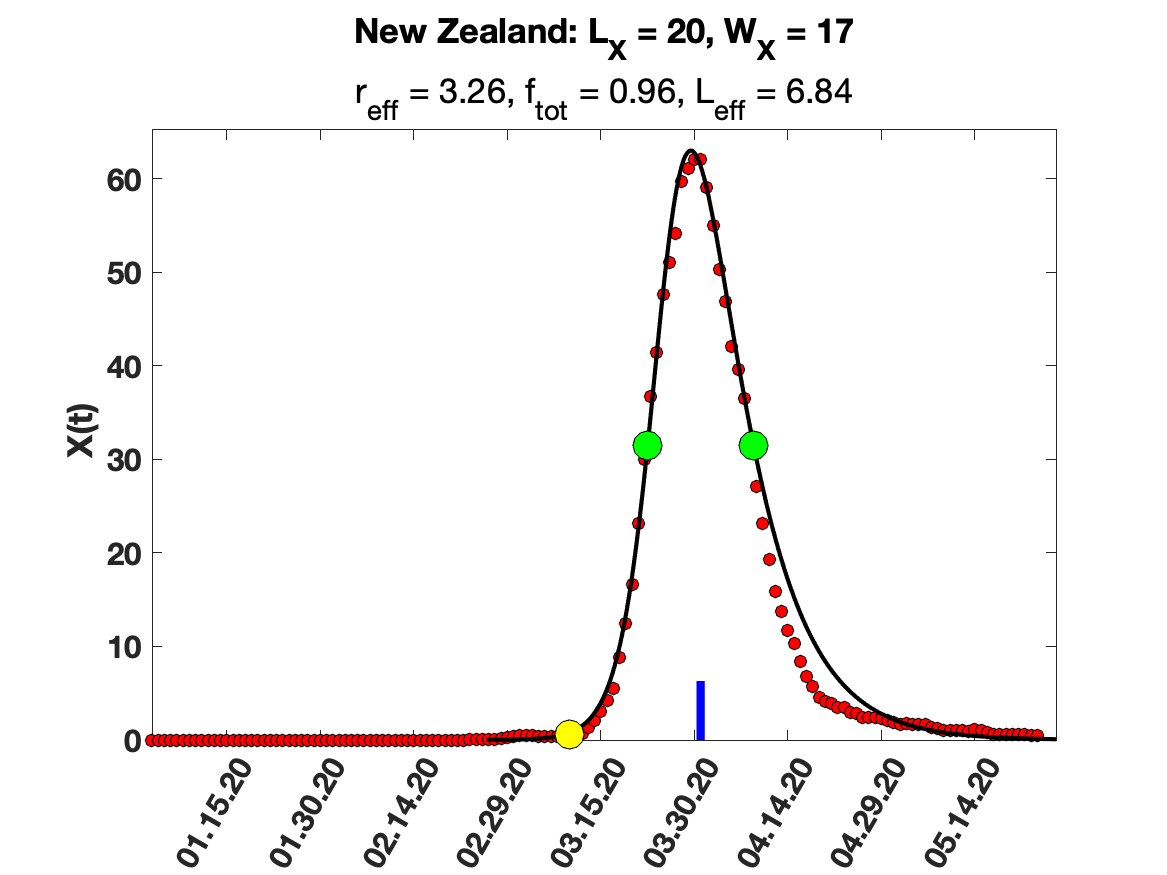

### Norway.png

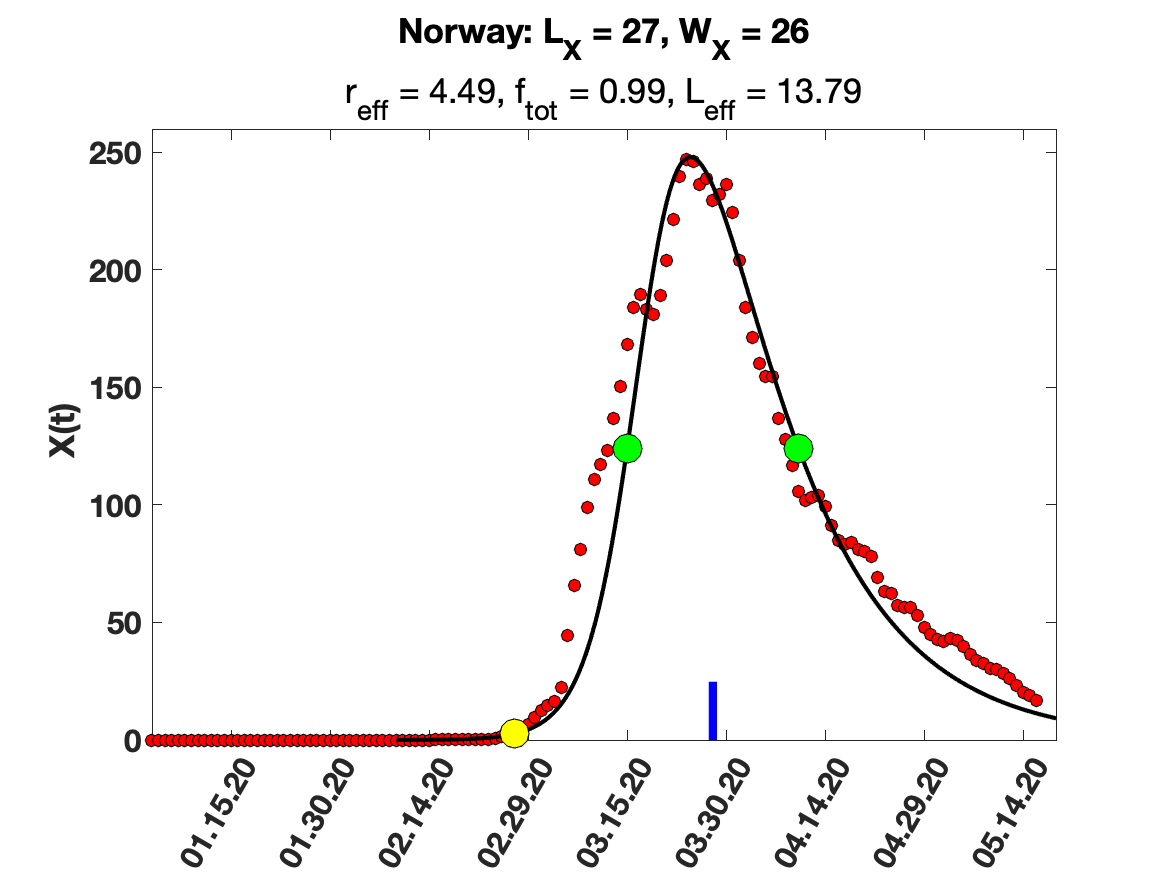

### Portugal.png

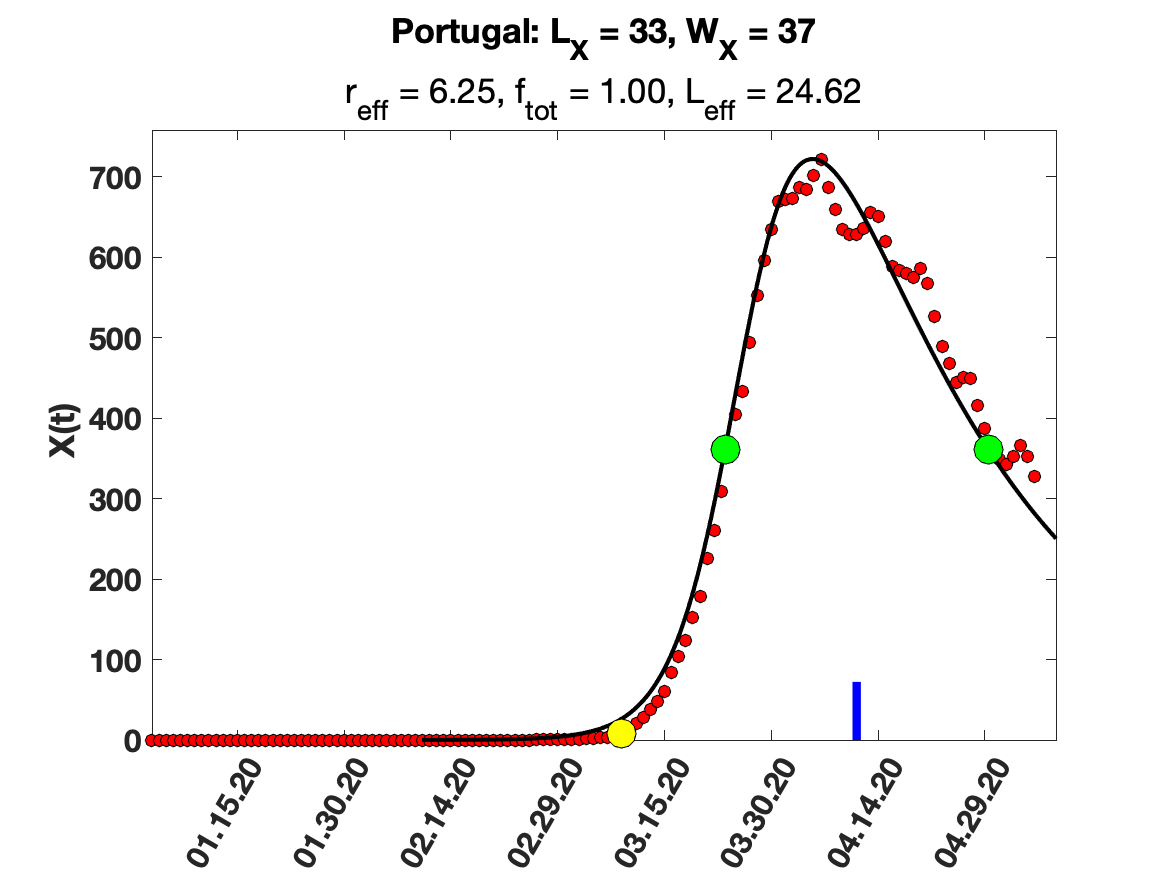

### Qatar.png

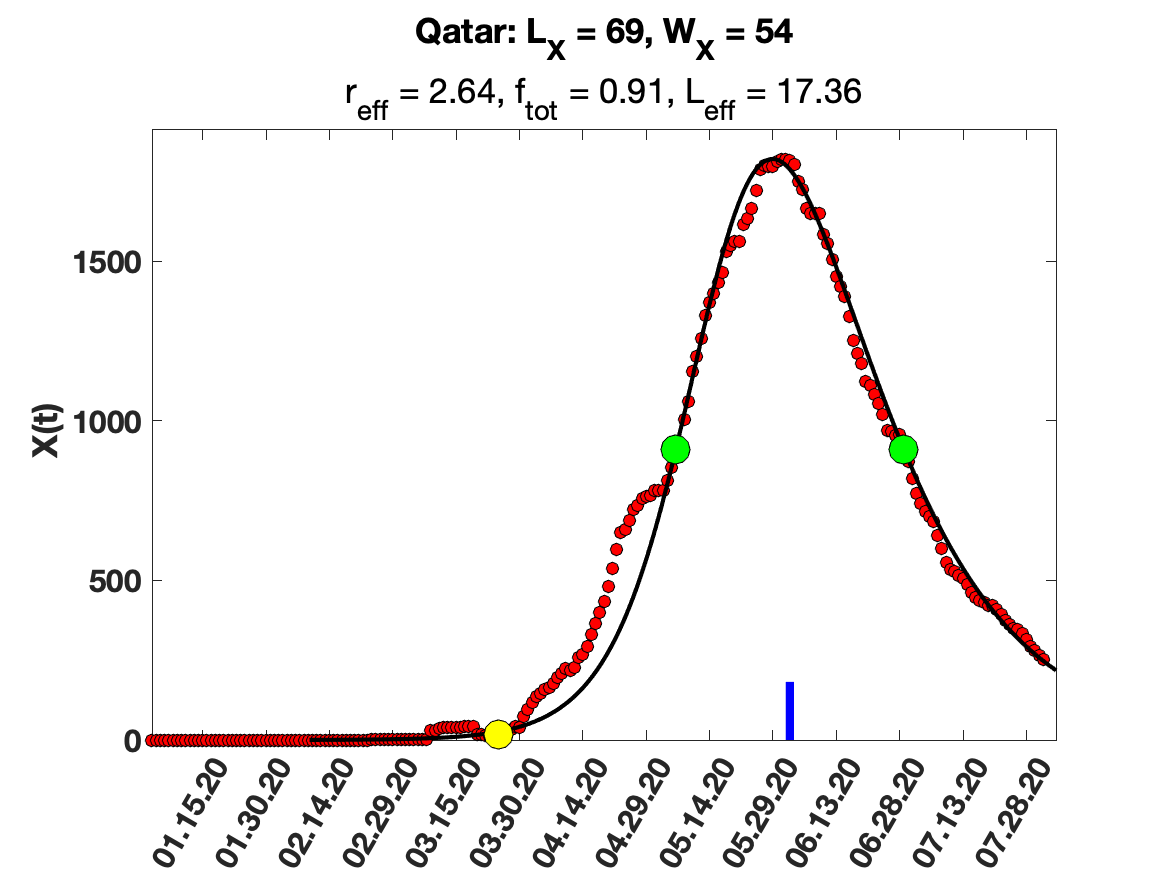

### Serbia.png

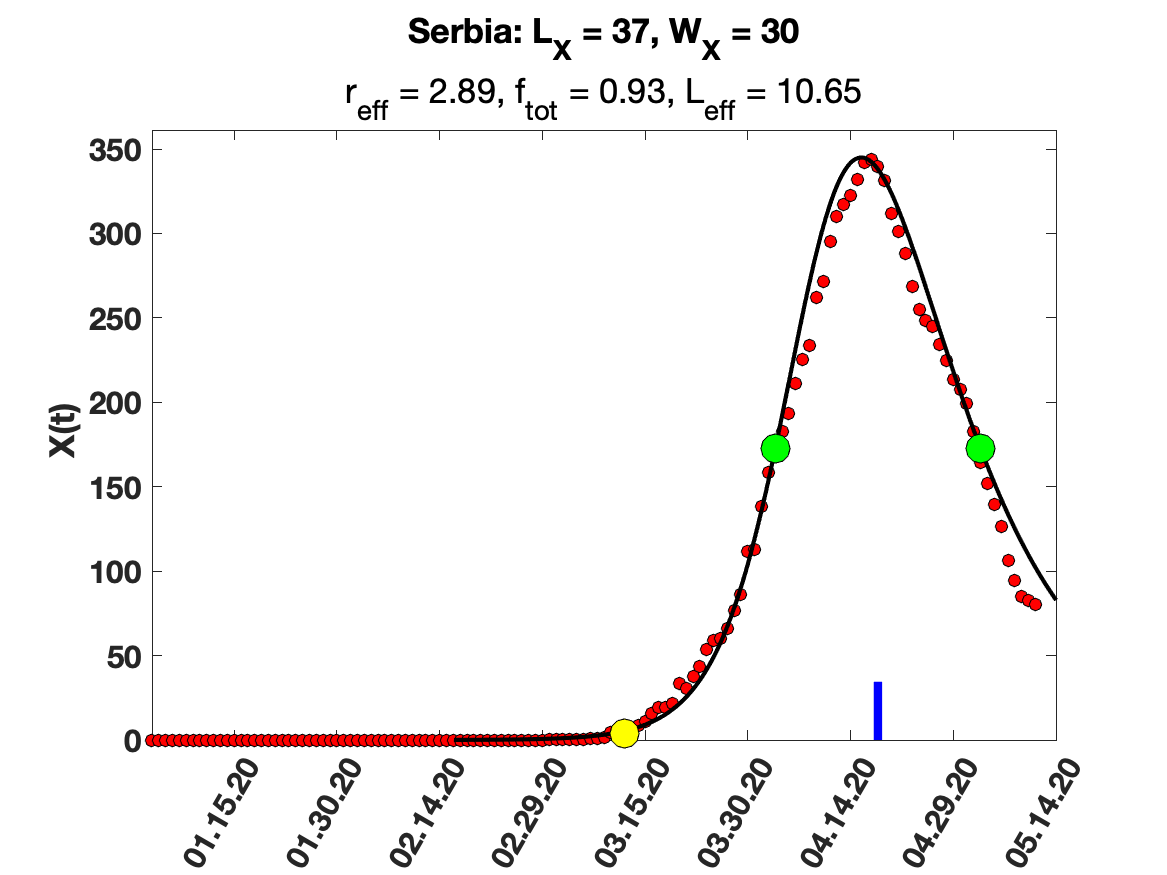

### South Africa.png

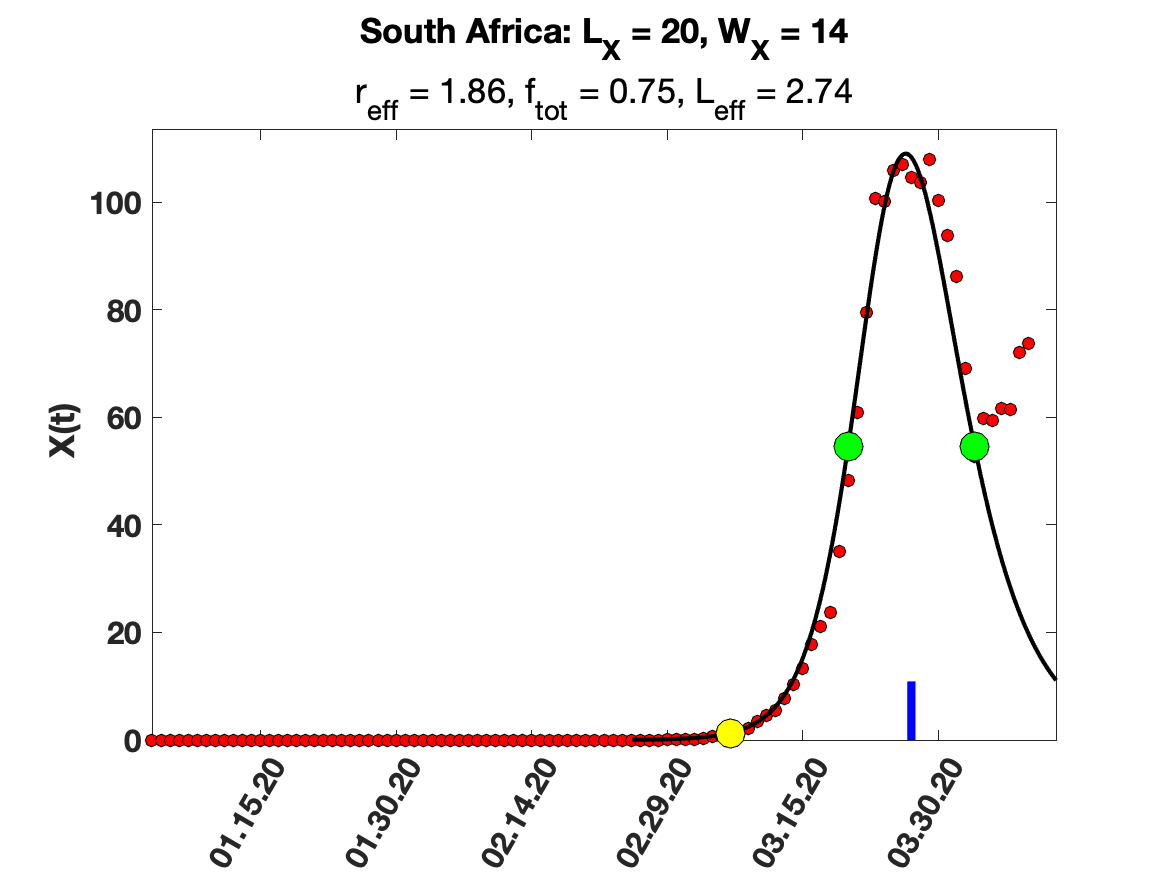

### South Korea.png

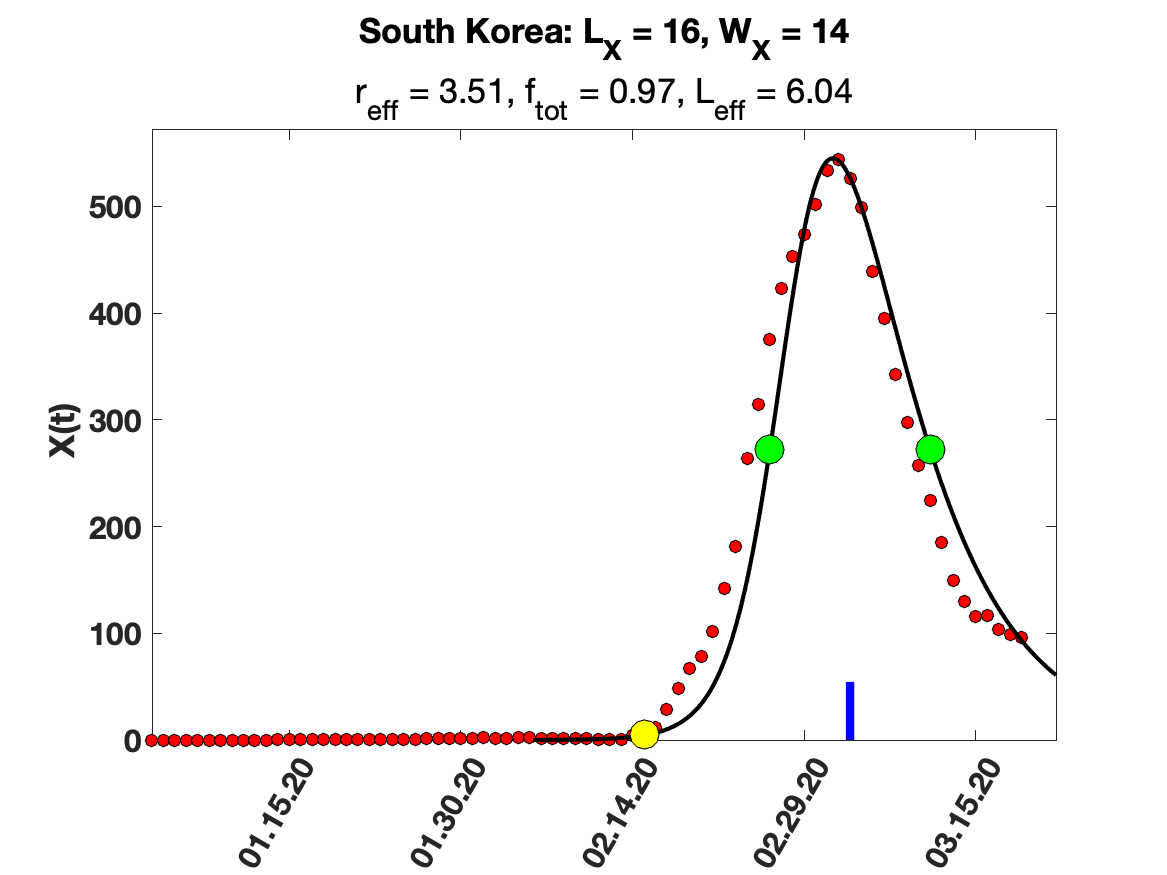

### Spain.png

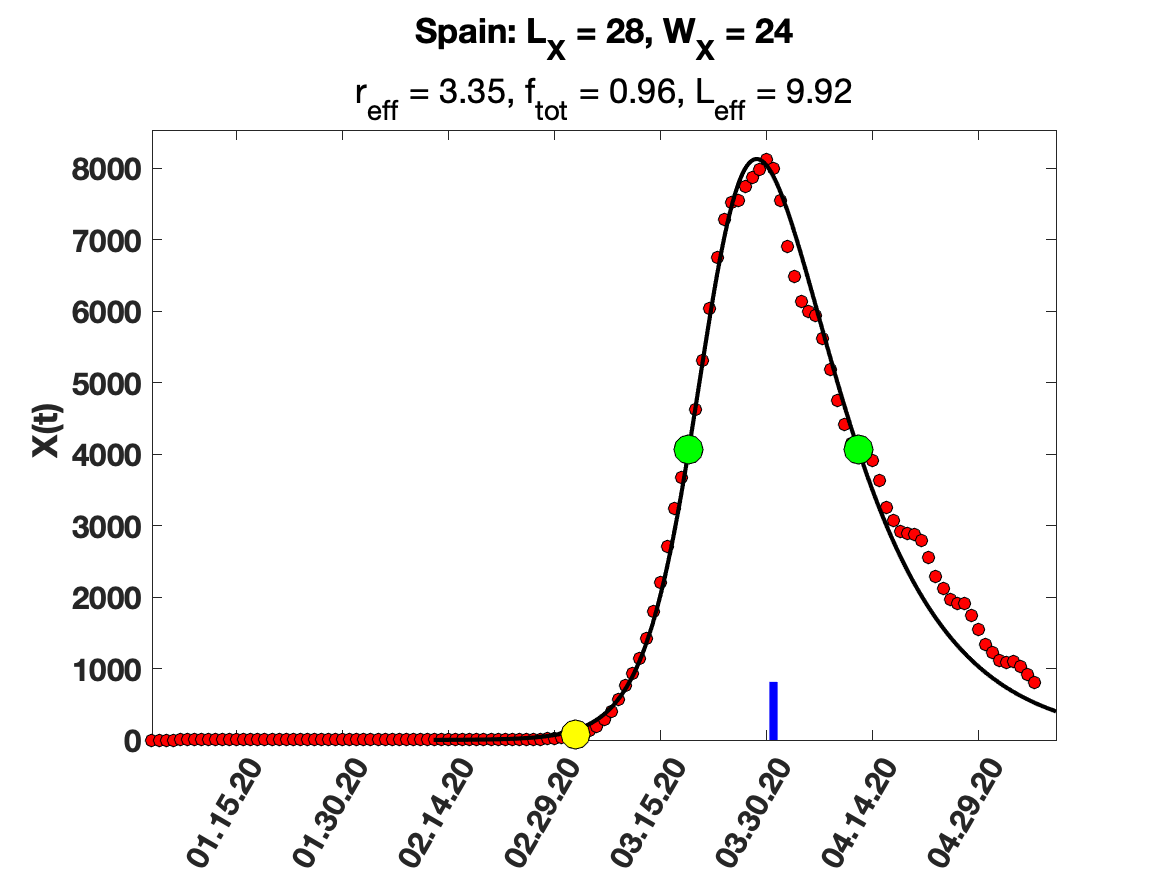

### Switzerland.png

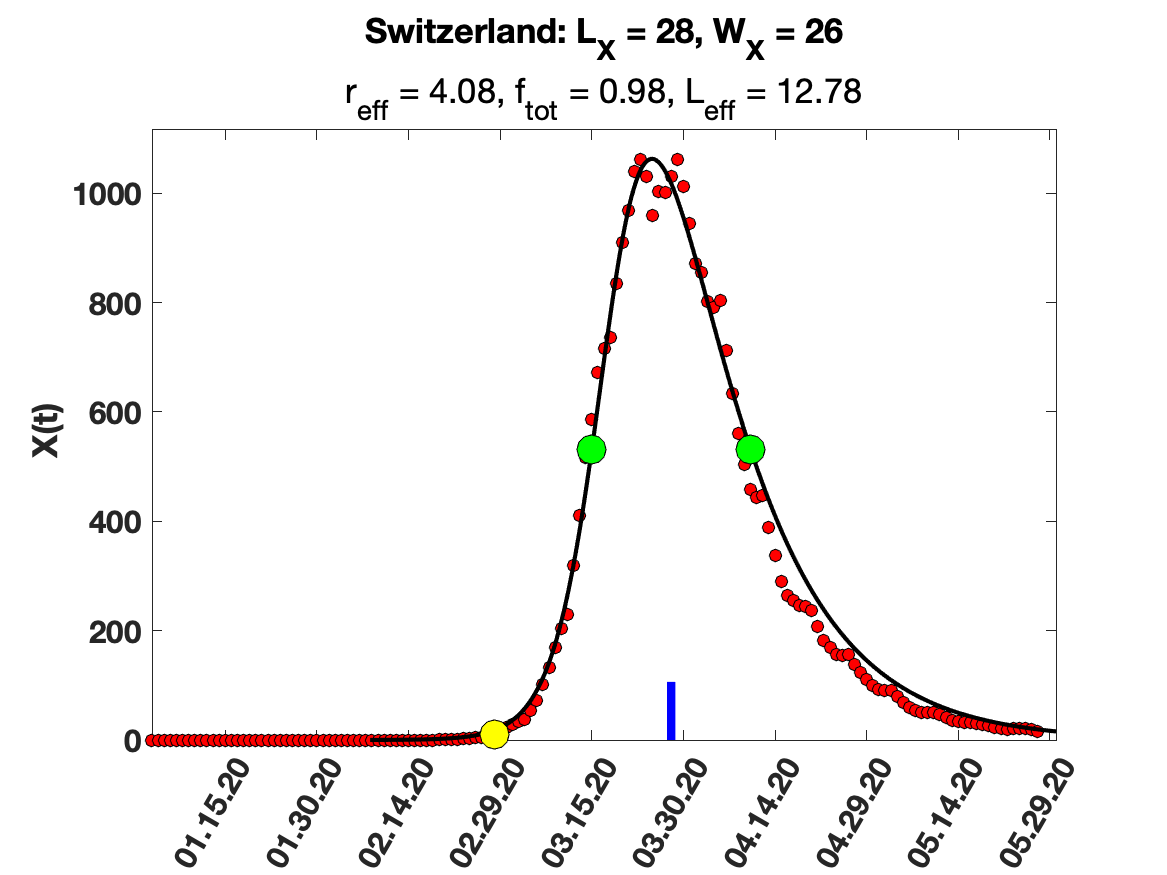

### Tajikistan.png

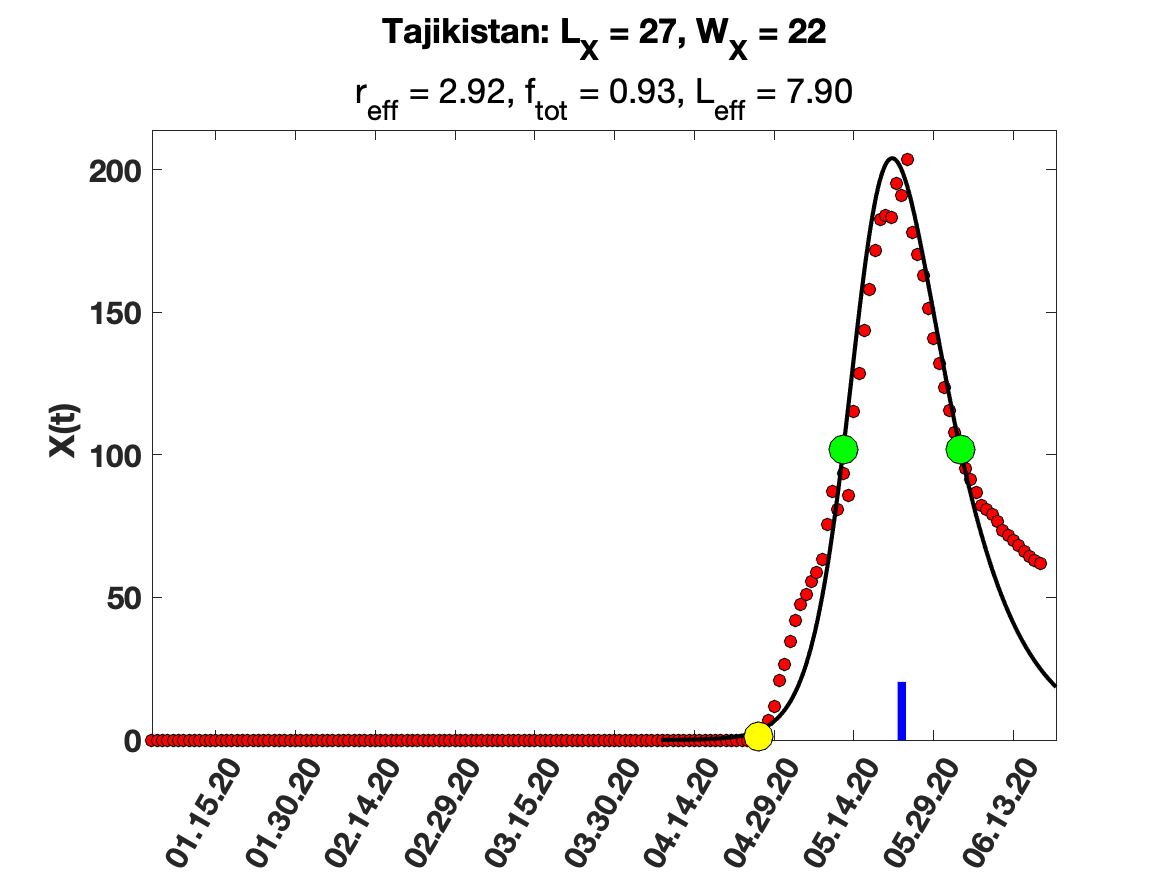

### Thailand.png

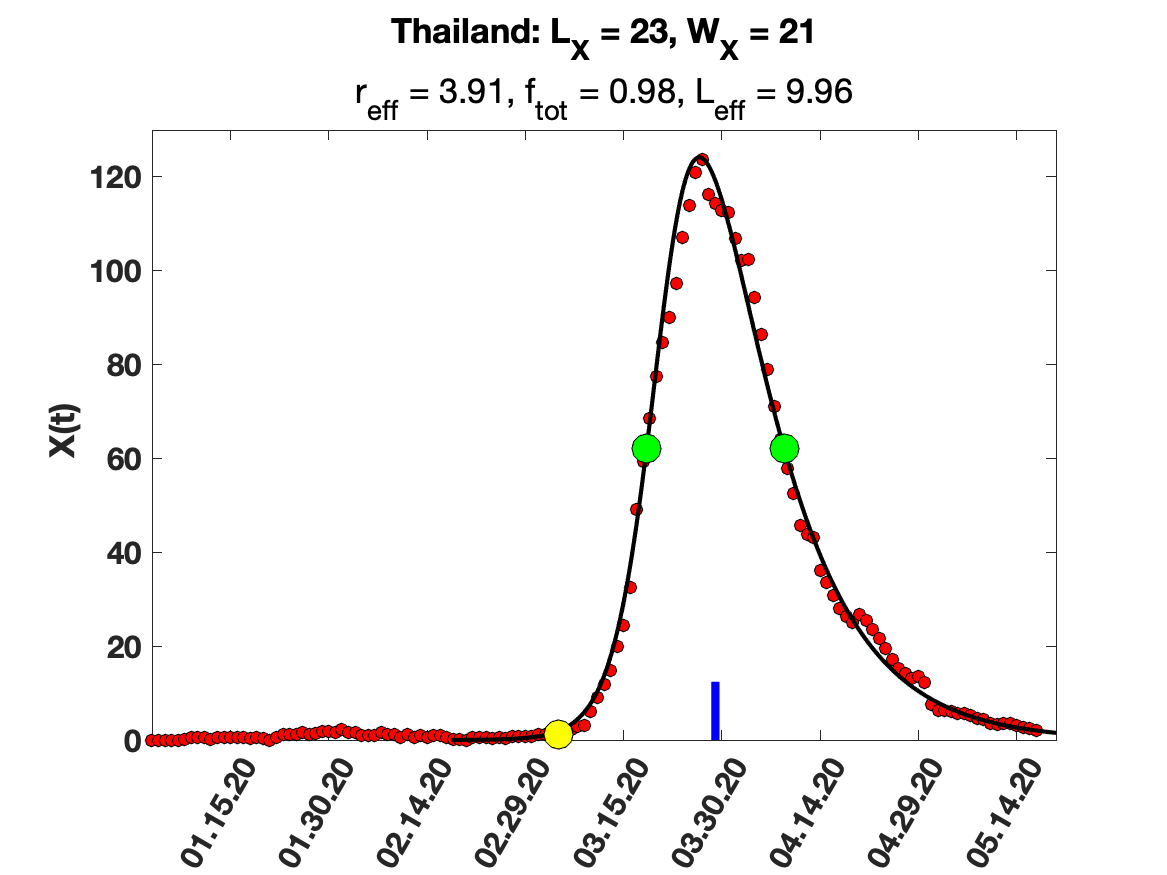

### The United Kingdom.png

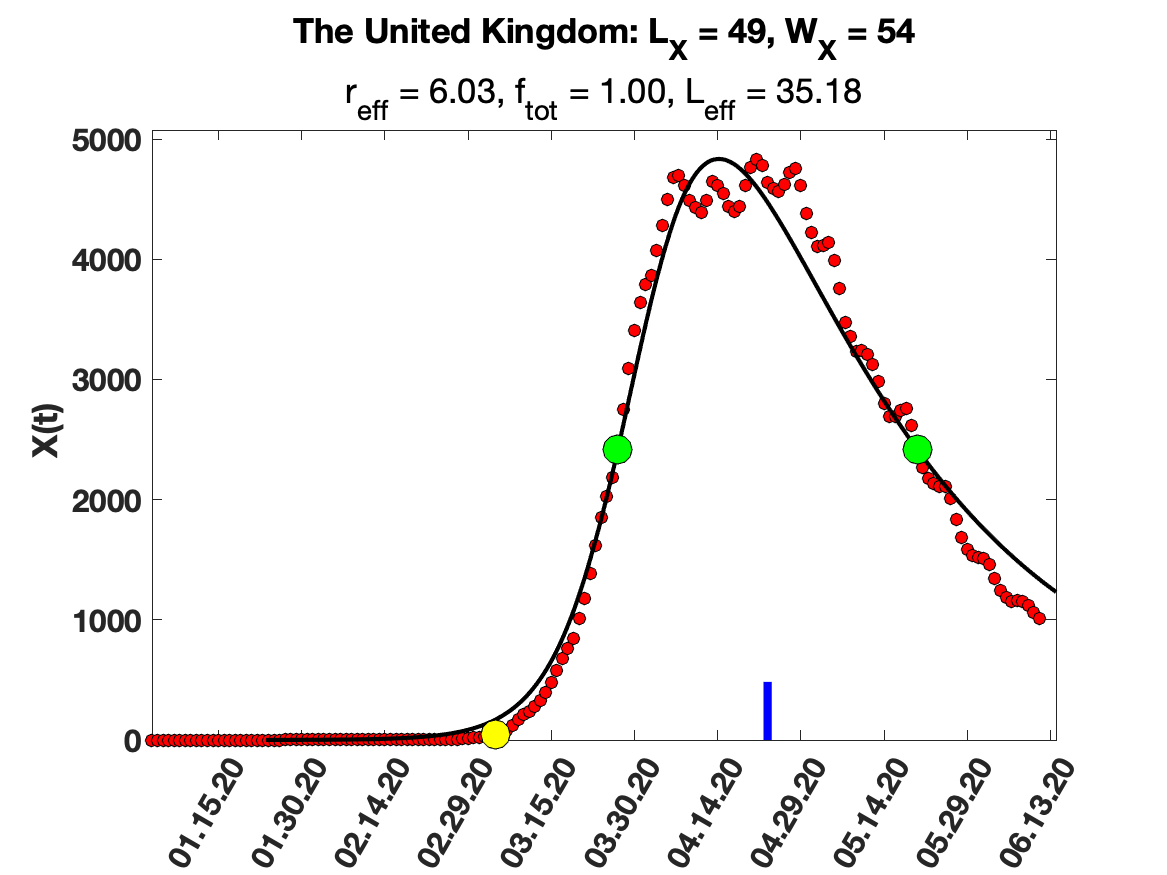

### Tu╠êrkiye.png

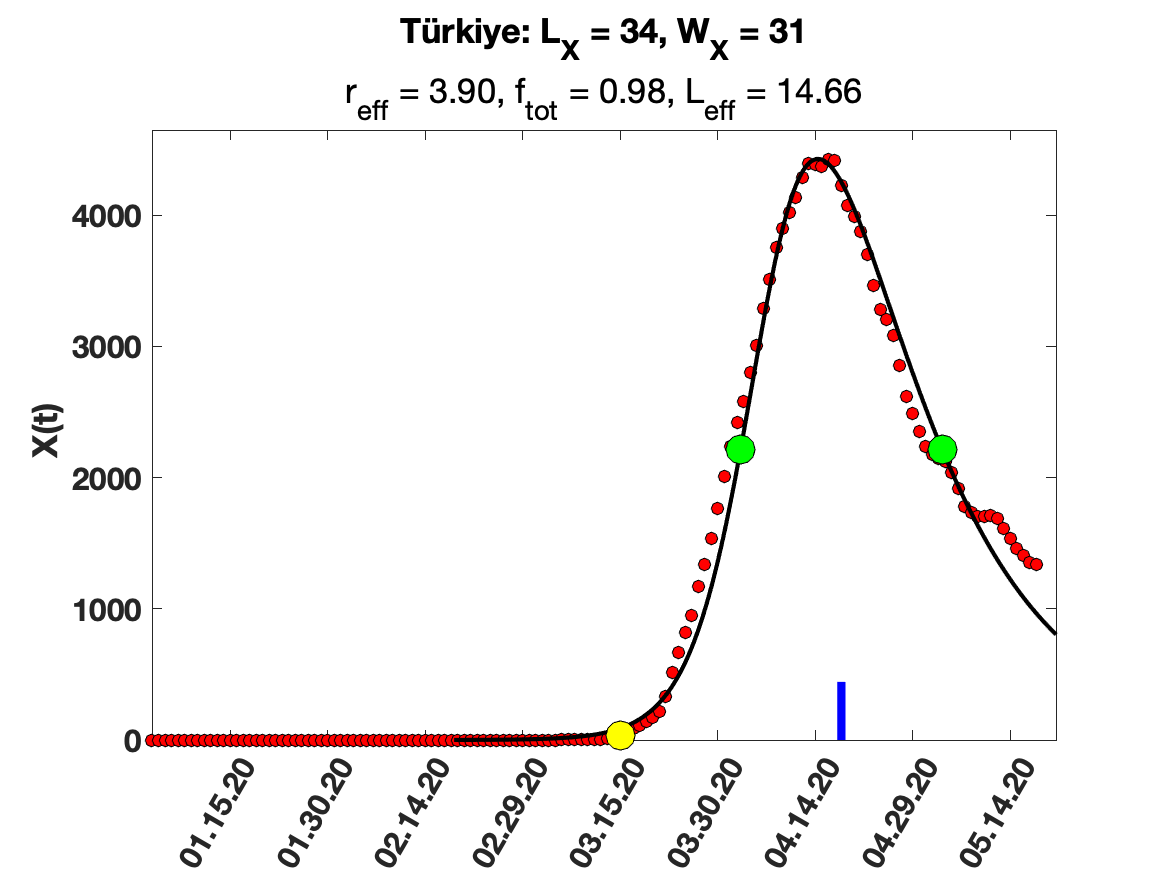
