## Appendix A for "Scaling rules for pandemics: Estimating infected fraction from identified cases for the SARS-Cov-2 Pandemic"

The rescaled equations for the pandemic dynamics are:

$$\frac{ds(\tau)}{d\tau} = -r_{eff}\, s(\tau)i(\tau) \qquad \text{(A1)}$$
$$\frac{di(\tau)}{d\tau} = r_{eff}\, s(\tau)i(\tau) - i\,(\tau) \qquad \text{(A2)}$$
$$\frac{dr(\tau)}{d\tau} = i(\tau) \qquad \text{(A3)}$$

The scaled quantities $s(\tau)$, $i(\tau)$ are related to S(t), I(t) of the SIR model by:

$s(\tau) = S(t)/N$ (A4)
$i(\tau) = I(t)/N$ (A5)
with, $\tau = \gamma_{eff} t = \frac{t}{L_{eff}}$ (A6)

Dividing (A2) by (A1) gives:

$$\frac{di(\tau)}{ds(\tau)} = \frac{1}{r_{eff}s} - 1 \qquad \text{(A6)}$$

Using the large N boundary conditions s(0) = 1, i(0) = 0 generates the exact result:

$$i(\tau) = 1 - s(\tau) + \log(s(\tau))/r_{eff} \qquad \text{(A7)}$$

At t=∞, $i(\tau) = 0$. Hence,

$$r_{eff}(s(\infty)) = -\frac{\log(s(\infty))}{1-s(\infty)} \qquad \text{(A8)}$$

When $s(\infty) = 1$ (no pandemic), l'Hospital's rule gives $r_{eff}(s(\infty) = 1) = 1$.

It is easy to see for $0 \leq s(\infty) < 1, r_{eff} > 1$. Hence, a pandemic requires $r_{eff} > 1$.

From (A2), the maximum in $i(\tau)$ happens when $s(\tau) = 1/r_{eff}$. Hence:

Maximum value of $i(\tau) = 1 - \frac{[1+\log(r_{eff})]}{r_{eff}}$, $r_{eff} > 1$ (A9)

Note that because of (A5) this quantity is the same as the maximum of I(t)/N which is the quantity $H_I/N$ in Eq. 12c in the main text. Hence,

$$H_I/N = 1 - \frac{[1+\log(r_{eff})]}{r_{eff}},\ r_{eff} > 1 \qquad \text{(A10)}$$

For small $\tau$, $s(\tau) \sim 1$. Hence, we can expand the right-hand side of (A7) in powers of (1- $s(\tau)$).

To lowest order,

$\log(s(\tau)) = \log[1 - (1 - s(\tau))] \cong -(1 - s(\tau))$ (A11)

Substituted into (A7) gives,

$\mathrm{i}(\tau) = \frac{\mathrm{r_{eff}}-1}{\mathrm{r_{eff}}}\left(1 - s(\tau)\right)$ (A12)

Substituting from (A12) into (A1) gives the Logistic Equation:

$\frac{\mathrm{d}s(\tau)}{\mathrm{d}\tau} = -(\mathrm{r_{eff}} - 1)s(\tau)(1 - s(\tau))$ (A13)

whose solution, with the boundary condition s(0) = 1-ε is:

$\mathrm{s}(\tau) = \frac{1}{[1+\varepsilon\, \mathrm{e}^{(\mathrm{r_{eff}}-1)\tau}]}$ (A14)

Hence, for $\tau \leq \frac{\log(\varepsilon)}{(1-\mathrm{r_{eff}})}$,

$\mathrm{s}(\tau) = [1 - \varepsilon\, \mathrm{e}^{(\mathrm{r_{eff}}-1)\tau}]$, (A15)

Combining (A12) and A(15) shows that for $\tau \leq \frac{\log(\varepsilon)}{(1-\mathrm{r_{eff}})}$,
$\mathrm{i}(\tau) = \frac{\mathrm{r_{eff}}-1}{\mathrm{r_{eff}}}\, \varepsilon\, \mathrm{e}^{(\mathrm{r_{eff}}-1)\tau}$ (A16a)

Hence, from Eq. 9,

$\mathrm{x}(\tau) = \omega\gamma_1 \mathrm{i}(\tau)/\gamma_{\mathrm{eff}} = (\frac{\omega\gamma_1}{\gamma_{\mathrm{eff}}})\, \frac{\mathrm{r_{eff}}-1}{\mathrm{r_{eff}}}\, \varepsilon\, \mathrm{e}^{(\mathrm{r_{eff}}-1)\tau}$ (A16b)

This shows that,

Exponent of in the initial exponential increase in cases for $\mathrm{x}(\tau) = (\mathrm{r_{eff}} - 1)\tau$ (A17a)
Exponent in the initial exponential increase in cases for $\mathrm{x(t)} = (\mathrm{r_{eff}} - 1)\gamma_{\mathrm{eff}}\, \mathrm{t}$ (A17b)
