## Appendix B for "Scaling rules for pandemics: Estimating infected fraction from identified cases for the SARS-Cov-2 Pandemic"

We will show an example of the use of the data in Supplementary Table 1 and Figure 1 a-f to find $f_{tot}$, $r_{eff}$, $L_{eff}$ and $\alpha$ using only data for X(t), the symptomatic/identified cases per day (Eq. 5). Consider a numerical solution of Eq. 1-5 for parameter values: $N = 10^4$, $L_0 = 10$ days, $L_1 = 5$ days, $L_{eff} = 8$ days, and $r_{eff} = 1.5$ which, from Eq. 4b and Eq. 10, corresponds to $\omega = \frac{\left(\frac{L_0}{L_{eff}} - 1\right)}{\left(\frac{L_0}{L_1} - 1\right)} = 0.25$ and $\alpha = 0.1875$ respectively. The functional form of S(t), I(t), R(t) and X(t) in Figs. B1a-d was obtained by numerically solving Eq. 1-5 using the stiff ODE solver ode15s in Matlab with boundary condition: $S(0) = 1- \varepsilon N$, $I(0) = \varepsilon N$ and $R(0) = 0$, with $\varepsilon = 0.001$. The result obtained are shown in Figs. B1 a-d. The measured total infected fraction was found to be $f_{tot} = 0.584$. To make contact with real data and to measure $L_X$, we need an objective definition of "the start day of the pandemic." We define this as the day when the number of recorded daily cases is 1% of the peak in daily cases, which was also the criterion used for the data in Supplementary Table 1.

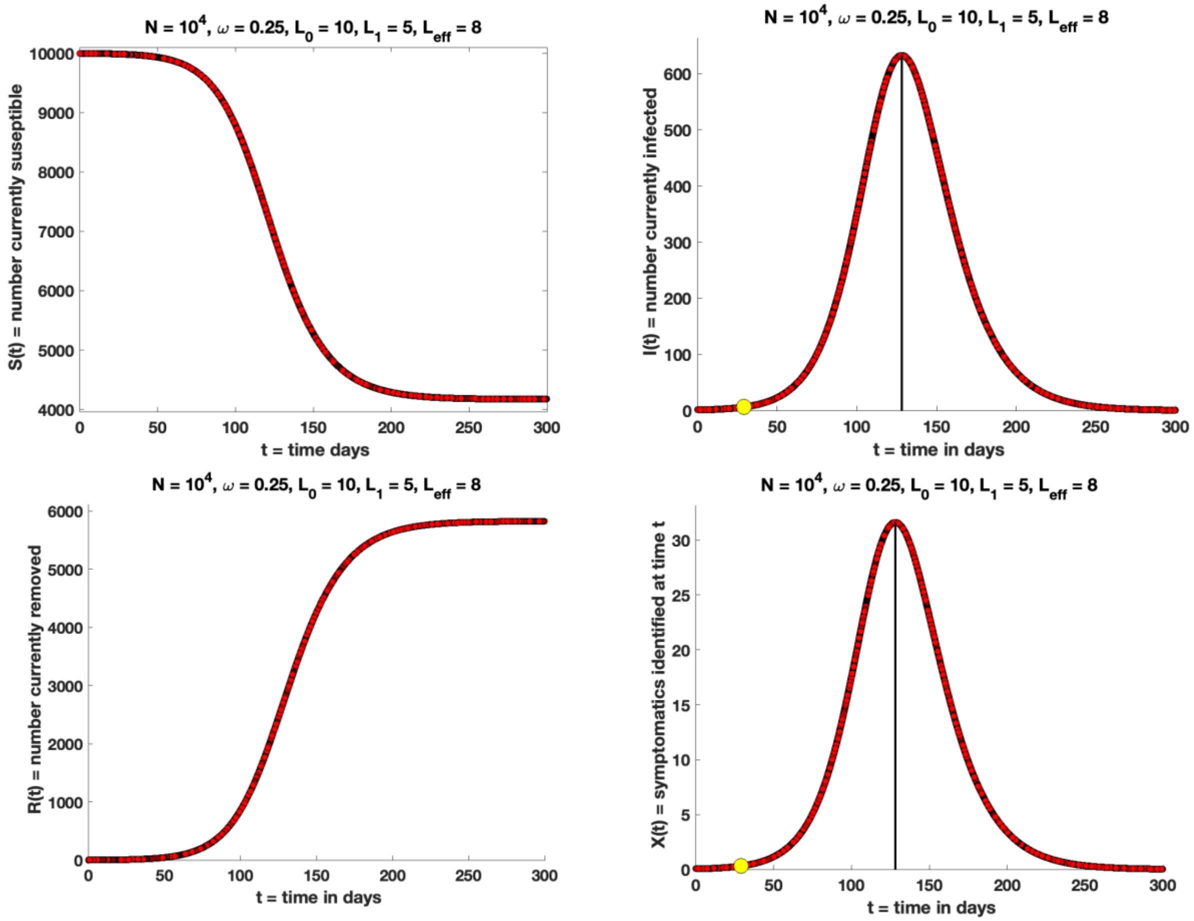


Fig. B1a-d: Solution of Eq 1-5 using the parameters shown. *Note that the yellow dots in Figs. B1b and B1d are our objective definitions of the "start of the pandemic," i.e., the day when the number of cases is 1% of the peak.* The same definition was used in generating the data in Supplementary Table 1. $L_X$ is the number of days from the yellow dot in Figure B1d to the location of the peak. From Fig. B1d, we find, $L_X = 98.9$ days, $W_X = 64.9$ days, and $f_{tot} = 0.58$.

Now imagine that we only know X(t) (Fig B1d but without the parameters in the title), and not the other data in Fig B1a-c (shown in Fig B1e). Using this information alone, we want to estimate $f_{tot}$, $r_{eff}$, $L_{eff}$ and α. From the data for X(t), we can find the location of the peak and $W_X$ = 64.87 days, the half width at full maximum. We can also find $L_X$ = 98.85 days from the number of days from when X(t) was 1% of the peak to the location of the peak.Thus $L_X/W_X$ = 1.53. Using this value in Supplementary Table 1 gives the correct values $f_{tot}$ =0.58 and $r_{eff}$ = 1.5. Using the values $L_X/L_{eff}$ = 12.37 and $W_X/L_{eff}$ = 8.10 for $r_{eff}$ = 1.5 in Supplementary Table 1, we get two estimated values 8.01 days and 7.99 days respectively for $L_{eff}$. Finally, we can estimate $\alpha = \frac{r_{eff}}{L_{eff}} = 0.1875$.

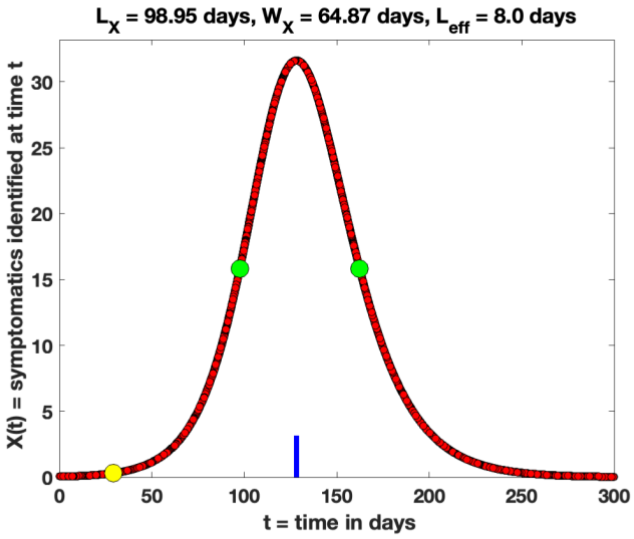


Fig. B1e: Inferring parameters from only X(t). The yellow dot defines the "start" of the pandemic, the day the number of cases is 1% of the peak. The same definition was used in generating the data in Supplementary Table 1. $L_X$ is the number of days from the yellow dot to the location of the maximum (shown as a blue mark) and $W_X$ is the half width of the peak in X(t) (the time between the green dots). This gives $L_X/W_X$ = 1.53, which from the data in Supplementary Table 1 gives $f_{tot}$ =0.58 and $r_{eff}$ = 1.5. Two estimates for $L_{eff}$ can be obtained from the data for $L_X/L_{eff}$ and $W_X/L_{eff}$ for this value of $r_{eff}$ in Supplementary Table 1.
